# Dear Watch, Should I Get a COVID-19 Test? Designing deployable machine learning for wearables

**DOI:** 10.1101/2021.05.11.21257052

**Authors:** Bret Nestor, Jaryd Hunter, Raghu Kainkaryam, Erik Drysdale, Jeffrey B. Inglis, Allison Shapiro, Sujay Nagaraj, Marzyeh Ghassemi, Luca Foschini, Anna Goldenberg

## Abstract

Commercial wearable devices are surfacing as an appealing mechanism to detect COVID-19 and potentially other public health threats, due to their widespread use. To assess the validity of wearable devices as population health screening tools, it is essential to evaluate predictive methodologies based on wearable devices by mimicking their real-world deployment. Several points must be addressed to transition from statistically significant differences between infected and uninfected cohorts to COVID-19 inferences on individuals. We demonstrate the strengths and shortcomings of existing approaches on a cohort of 32, 198 individuals who experience influenza like illness (ILI), 204 of which report testing positive for COVID-19. We show that, despite commonly made design mistakes resulting in overestimation of performance, when properly designed wearables can be effectively used as a part of the detection pipeline. For example, knowing the week of year, combined with naive randomised test set generation leads to substantial overestimation of COVID-19 classification performance at 0.73 AUROC. However, an average AUROC of only 0.55 ± 0.02 would be attainable in a simulation of real-world deployment, due to the shifting prevalence of COVID-19 and non-COVID-19 ILI to trigger further testing. In this work we show how to train a machine learning model to differentiate ILI days from healthy days, followed by a survey to differentiate COVID-19 from influenza and unspecified ILI based on symptoms. In a forthcoming week, models can expect a sensitivity of 0.50 (0-0.74, 95% CI), while utilising the wearable device to reduce the burden of surveys by 35%. The corresponding false positive rate is 0.22 (0.02-0.47, 95% CI). In the future, serious consideration must be given to the design, evaluation, and reporting of wearable device interventions if they are to be relied upon as part of frequent COVID-19 or other public health threat testing infrastructures.

## INTRODUCTION

The use of wearable devices has skyrocketed in recent years - about 3 in 10 people in the USA wear a fitness or health tracking sensor [62]. The abundance in physiological data has lead to the development of algorithms to deliver healthcare at the edge, with regulatory approval in applications such as atrial fibrillation [45]. In the wake of the global COVID-19 pandemic there has been unprecedented need for early detection of COVID-19 in a way which differentiates it from seasonal influenza or otherwise unspecified influenza-like-illnesses (ILI), due to the substantial overlap in signs and symptoms. Jurisdictions have circumvented transmission through extensive testing and proper infection control through physical distancing, isolation of positive cases, and contact tracing [26; 48]. In contrast, a reduced capacity for testing has been identified as a rate-limiting step in the control of the pandemic [25]. Wearable devices can contribute to testing infrastructure, and in turn, reduce transmission. Studies conducted prior to and throughout the pandemic demonstrate that the physical manifestations of ILI, and in particular, COVID-19 can be captured using wearable devices [35; 38; 43; 51]. To understand the usefulness of wearable devices for the public health infrastructure, proposed models must be evaluated in a scenario that resembles conditions of practical deployment. It is essential to exhaustively evaluate the potential impacts of models based on wearable data to avoid unattainable expectations or under-utilisation.

Current testing infrastructure is typically limited to COVID-19 RT-PCR tests^1^, which are invasive and only show a single time-point that determines whether the viral load of SARS-COV2 is above a detectable threshold. The properties of PCR are such that they are highly specific (i.e. COVID-19 negative patients are hardly detected as COVID-19 positive 98%) and reasonably sensitive (71-98% of tests on COVID-19 positive individuals are positive [57; 59; 61]). The highest viral load is detected on the days surrounding the onset of symptoms [60]. However, many individuals are only encouraged to get a COVID-19 test if they have had known contact with a COVID-19 case, or have already shown symptoms. It is estimated that 44% of viral transmission occurs prior to the onset of symptoms in cases where symptoms eventually develop [17]. This is problematic as false negative rates from the test are elevated prior to symptom onset [27]. Emerging evidence for COVID-19 estimates that 17.9% of cases are asymptomatic [39]. As with influenza, asymptomatic COVID-19 cases can still transmit the virus [30], though their transmission may be at lower odds than symptomatic individuals [4].

Accessible and frequent COVID-19 tests, even if at lower sensitivity, have the potential to undercut viral transmission if SARS-COV2 harbouring individuals are notified during or prior to peak infectiousness. Simulations have shown that by using frequent, low-sensitivity COVID-19 tests it is possible to rapidly detect and contain outbreaks of the virus on university campuses [44]. Simulated daily tests with sensitivities as low as 70% (and 98% specificity) result in 162 cases out of a cohort of 4990 over an 80 day semester [44]; this is in stark contrast with symptom based screening estimates 4970 of 4990 would be infected, although this assumes only 30% of infected individuals show symptoms and the reproductive number, *R*_*t*_, is 2.5. It is presumed that many low viral-RNA-count, positive-PCR tests are beyond peak infectiousness (possibly even no longer infectious) due to the long RNA-positive tail [36; 37]. This emphasises the need for early testing. Using frequent tests simulates better control of the pandemic than improving the sensitivity of tests [28]. While it is possible to acquire and administer rapid tests, we should utilise the infrastructure already available, including the sensors on our wrists which can passively monitor for ILI events.

Unfortunately, there is no sensor for measuring SARS-CoV-2 viral load in consumer wearable devices that are widely used. Instead we must rely upon the physical manifestations of the infection to trigger a classification. Fever is a hallmark symptom of infection that is characterized by marked elevation in basal body temperature. While some consumer wearables are capable of measuring temperature [55], many are primarily intended as fitness related devices equipped with optical heart rate sensors and accelerometers. Despite the lack of direct temperature monitoring, body temperature is mediated by and affects cardiac rhythm and function [24]. In influenza, evidence indicates that there is an increase in resting heart rate (RHR) correlated with onset of fever [14]. Abnormally high heart rate measurements from wrist based devices have been associated with inflammatory illnesses [33]. Increases in RHR, impaired sleep due to illness, and reduced steps due to inactivity measured using Fitbit user data have been correlated with influenza rates at a population-level [51]. Other available wearable measurements, such as heart rate variability (HRV) and respiratory rate, also correlate to infection [35; 43].

Using wearable devices as diagnostic tools is enticing, but to truly understand their potential for this purpose, models that use wearable data must be assessed in frameworks similar to the deployment conditions and subject to the same scrutiny that any non-wearable-based diagnostic test would face. There are numerous examples in the literature outlining the prerequisites needed to build diagnostic models for wearable data [18] and yet the recently published papers [35; 38; 43] lack in one or another aspect as elaborated below.

In this work we address the design flaws in previous works’ evaluation methods which have so far lead to the overestimation of COVID-19 detection; this is done by intentionally designing the study so that we can estimate post-deployment performance. First, model performance is compared in the format of current literature. We highlight the strengths and weaknesses of these reporting paradigms. After, we demonstrate the actual performance one can expect in a forthcoming week for COVID-19 detection using wearable devices only, survey data only, and a combination of wearable data and survey data.

While we are able to obtain similar performance to those published under their evaluation procedures, in a realistic framework the wearable contribution is likely to be much weaker. Despite lower performance, implementation of the wearable device prediction models results in a reduction in the number of symptom-based surveys required to detect COVID-19 cases, and elevates the likelihood that a tested individual has COVID-19 compared to a randomly tested sample of the population. Maximizing the pre-test probability of COVID19 ensures that we do not over-test, an important consideration for public health.

## RESULTS

We collected a dataset, referred to as `FLUCOVID`, which contains survey and wearable device data collected between November 1, 2019, and June 1, 2020. There are 32, 198 participants with *both* surveys and wearable data in this cohort, 2, 554 had medically diagnosed influenza, 204 had COVID-19, and the remainder had unspecified ILI. A complete description of the data collection process can be found in [53]. Our survey data proportionally matched peaks and trends in CDC data for influenza [8] and COVID-19 incidences [16] (See Figure 1). Using self reported symptom onset and recovery dates, combined with reports of medically diagnosed COVID-19 we define the task as a daily classification of whether a date is within the self reported ILI symptom dates (label of 1) or outside of that region during presumably healthy days (label of 0). Under this paradigm, day 0 is the first day in which a participant experiences symptoms, despite acquiring the virus and still being capable of transmitting the virus prior to symptom onset (negatively labelled days in the range of -14 to -1).

**Figure 1.**
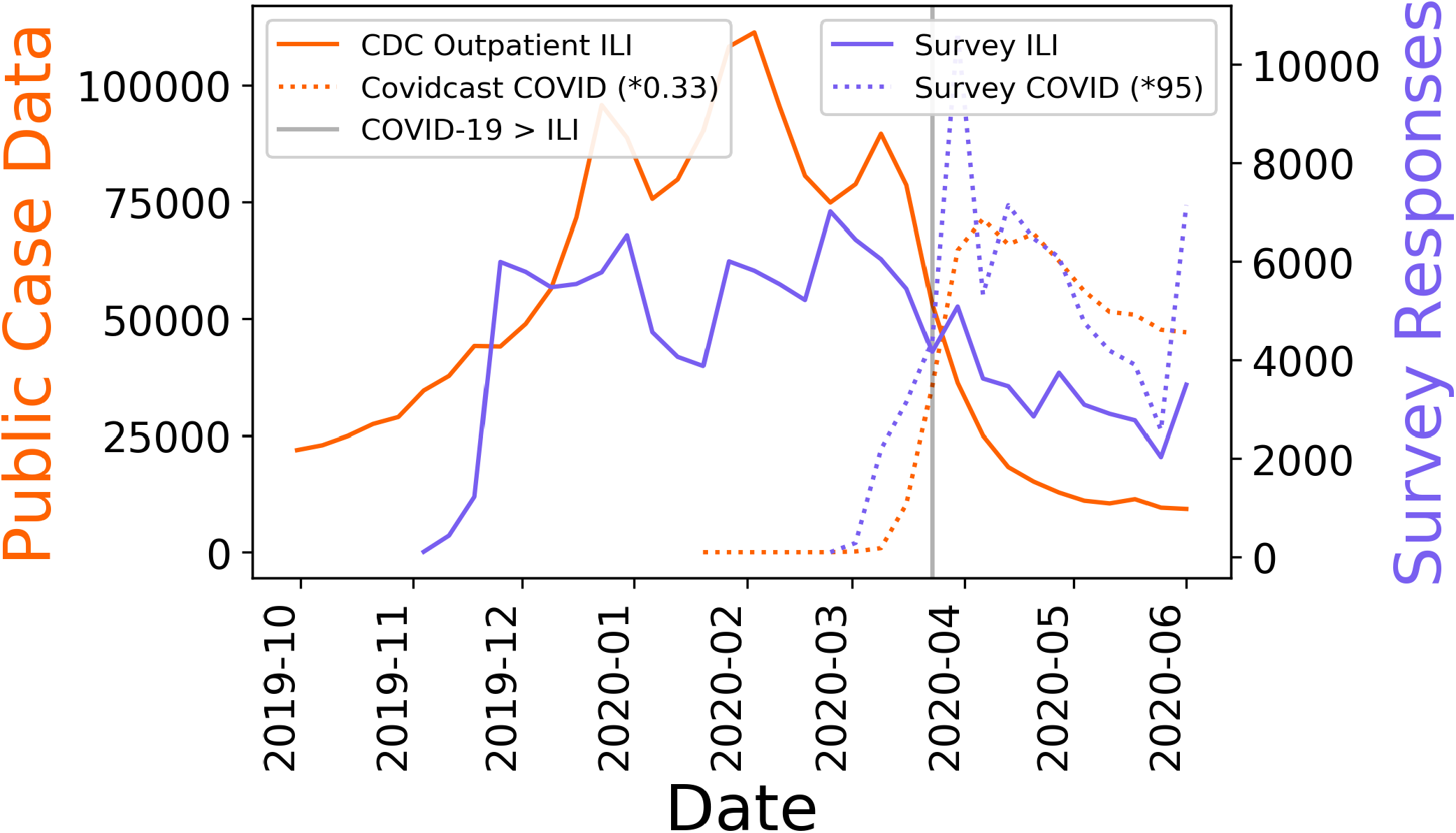
Influenza and COVID-19 incidence: Publicly available data for outpatient influenza from the CDC ILINet and COVIDcast case counts are compared to our incidence of survey ILI and COVID-19 cases. The COVID-19 data is scaled as denoted in the legend. After COVID-19 becomes more prevalent than ILI, as denoted by the vertical line, the ILI survey cases show an unusually high proportion of unspecified ILI, compared to the CDC outpatient counts. Some unspecified ILI after March 16 may be attributed to undiagnosed COVID-19.

We investigated several prepossessing steps for the input data and the outcome. We investigated normalizing individuals’ covariates by their own history as was previously done in the literature [51]. However, we found that this step did not outperform simple normalisation of the individual’s covariates by the training set population means. We thus normalise our data by the training set population means. This step allows to make more predictions immediately after a user enrols rather than waiting to accumulate sufficient data. In the spirit of using the most data available, we did not eliminate users with vast amounts of missing data due to lack of wear-time or missing channels. This deviates from the convention accepted in previous works [35; 38; 43; 50] but allows us to make predictions possible even in tough edge cases. To address the increased amount of missing data due to the broader inclusion criteria, we mean imputed missing data at the beginning of the study to left align the participants and forward filled data after an individuals’ steps, heart rate, or sleep time has already been observed. Note the XGBoost [12] and GRU-D [11] models are capable of handling missing data. See further details on input data pre-processing in the Online Methods and the Appendix.

For the survey response covariates, survey responses are given once per week recounting symptoms experienced on each day. These responses are disambiguated into binary daily symptom flags for each of the reported symptoms or behaviours. Unfortunately, some participants respond to surveys weeks after their ILI event to provide onset and recovery dates. These individuals are retained during all training and testing procedures. Wherever no information is reported it is assumed that participants are not experiencing any symptoms and are following all COVID-19 precautions. The day-level ILI labels are constructed by labelling all days between self-reported symptom onset and self-reported recovery as positive. This differs from previous work where a fixed number of days around symptom onset are labelled as positive days [35; 43; 50]. We did this in order to be able to make predictions continuously on any date prior to, or after symptom onset, to better simulate deployment. It is important to remember that our detector is not a wearable COVID-19 specific detector, it is a detector of ILI events that are a superset of any medically undiagnosed ILI, self-reported medically diagnosed flu, or self-reported medically diagnosed COVID-19. Finally, approximately a third of users had multiple reported events. When these events are separated by fewer than 7 days, they are merged into a single event, with the COVID-19 label superseding the unspecified ILI label.

All presented wearable models are trained using 48 variables representing sleep, heart rate, and steps measurements taken from wearable devices aggregated on a daily basis.

Throughout this work we use the following definitions to distinguish training and evaluation schemes which we compare in this paper:

### 1. Random splits

of data refers to a random shuffling of historic data to create train, validation, and test event splits. We also refer to it as **retrospective** evaluation, since the training events and testing events are all occurring in the past. In such a scenario, the prevalence of COVID-19 and ILI are likely to be the same across train and test, a conventional but not a realistic deployment scenario.

### 2. Prospective evaluation

is the setting where test data is chronologically after any of the training and validation datasets and data splits. We know that over a single season of data, influenza cases surge and fall, and with emergent COVID-19 infections cases are anything but stationary. The prevalence in the week after a COVID-19 wearable model deployment is unlikely to match the prevalence experienced in the training set when real life is concerned. Inevitably, a deployment-ready model would have to face this modelling challenge so it makes sense to test in this manner to avoid providing the public with poorly calibrated COVID-19 or ILI predictions.

### Aggregate metrics from randomly sampled cohorts artificially inflate performance

In the instance of seasonal influenza, models are able to use trends over time estimate current trends [51]. With emergent diseases like COVID-19, there is a lack of past season data to rely on to create such estimates. We trained XGBoost [12] model on 5 random splits of data (5 repetitions of 35% training, 7.5% validation, 7.5% held out retrospective test set, and 50% held out prospective test set by participant) to detect ILI events. When analyzing the performance of randomised test splits, we found that most of the AUROC metric was driven by between week comparisons, giving an inflated estimate of performance. For example, knowledge that first week of February had a high prevalence of influenza would lead the model to make more calls in the first week of February in the test set. Dissecting the test set predictions to their constituent weeks lowered the AUROC for nearly all weeks (See Figure 2). Removing the `week-of-year` feature helped mitigate the overestimate for the XGBoost model, but there was still performance drop from the overall AUROC, to the weekly AUROC on the randomly drawn test set (Figure 2). These results indicate that the model had learned to predict the prevalence of ILI in each week, rather than connecting the underlying physiological data to the outcome. Note that in each instance, the machine learning model outperforms a non-machine learning baseline that was designed for population level influenza surveillance [51].

**Figure 2.**
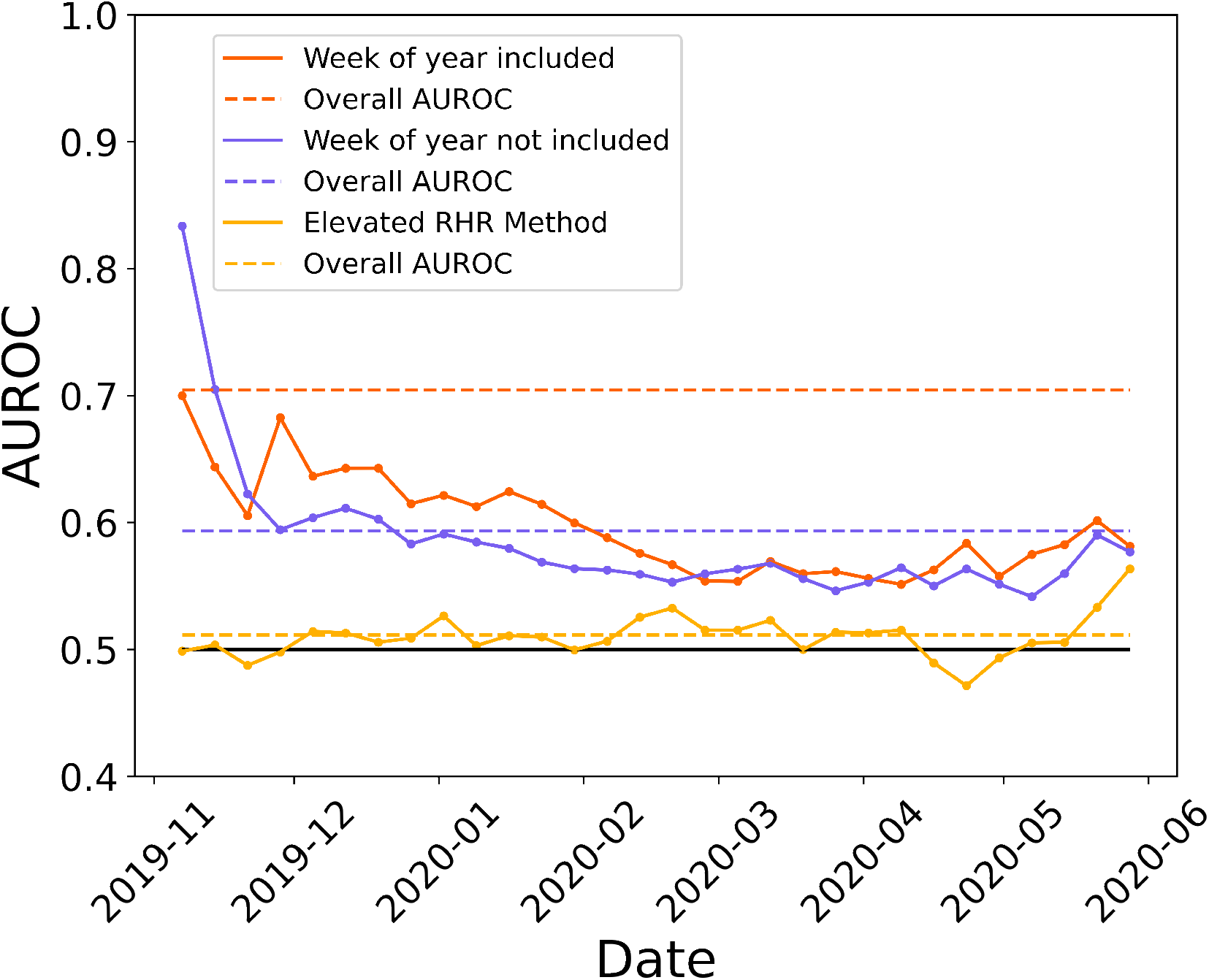
XGBoost trained on randomly split ILI data: Conditioning the performance by time (solid lines) reduces performance compared to when performance is assessed irrespective of time (dashed lines) for randomly drawn training, validation, and testing sets. All machine learning models outperform the non-machine learning elevated RHR baseline.

### Random retrospective test cohorts overestimate future performance

In emergent diseases, one would imagine training a model on all available data, selecting parameters, then deploying the model. The new cases that this model predicts on will be temporally disjoint from the training data. Then in the following week, the process would be repeated by training a freshly initialised model with an additional week of data. Naturally this creates a situation where there is a continuously growing training set, a sliding validation set window, and a sliding test set window (See Figure S1). This deployment-style evaluation is in contrast to the conventional ML **random split** dataset scenario used in the previous section and related work [43], since none of the future observations are seen during training. Additionally, our **prospective evaluation** only contains participants who are distinct from the participants in the training data for that week’s model for `FLUCOVID` data. Each consecutive week features a new, 50% of that week’s participants (sampled to obtain uncertainty estimates).

With each new model trained on the additional week of data, we also reserve a 7.5% **random split** of the data that is concurrent with the training set, since by the time of deployment, the participants in this set have already been encountered. This allows us to evaluate our models on a standard **random split** setting as one would in a typical ML workflow that is negligent to temporal patterns in data. We find that the perceived performance from the **random split** overestimates the **prospective evaluation** in nearly every week for both models (See Figure 3). Random test set evaluation overestimates our expected performance for ILI detection using wearable devices. These findings exemplify that the danger of overoptimistic result estimation is real when test setting is not representative of deployment scenario.

**Figure 3.**
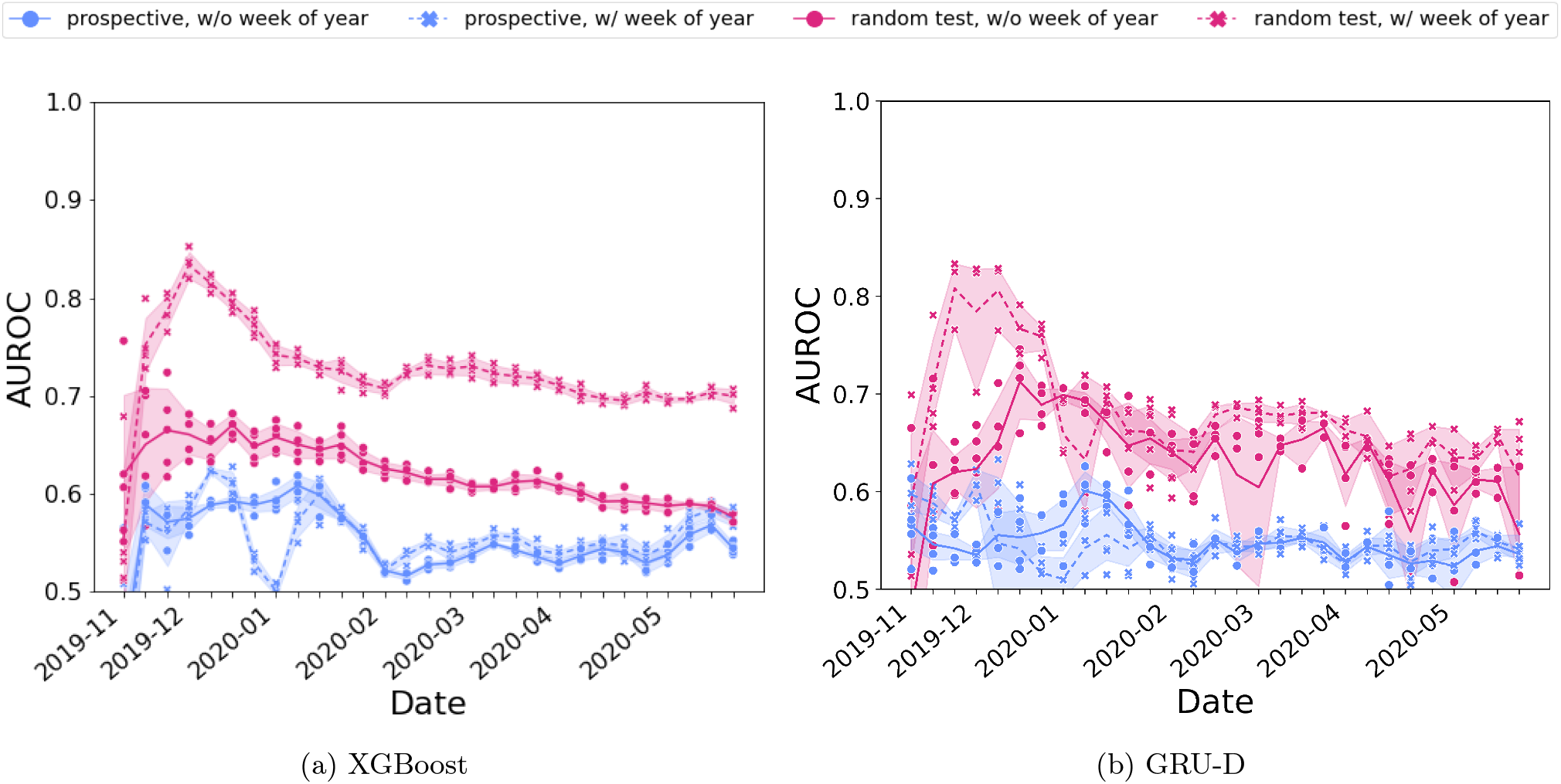
Random vs. prospective performance : Random retrospective test set AUROC compared to first week prospective test set AUROC for detection of ILI symptom days. Each week uses a newly initialised model trained on a random split of participants (50% prospective test set, 35% retrospective training set, 7.5% retrospective, last-week validation set, 7.5% retrospective test set split by participant).

Given the incidences of ILI and COVID-19 are non-stationary throughout the year, we attribute the drop in performance to an over-reliance on memorising the COVID-19 prevalence in a given week as before. This attribution is supported by the observation that by removing the `week-of-year` feature there is a corresponding drop in performance in retrospective test sets for XGBoost, which is not observed in prospective test sets (See Figure 3). To elicit the cause of the dataset shift, we apply PCA and t-SNE visualisations to the input data. When the `week-of-year` feature is absent, data do not show coherent clustering when falsely coloured by time. This temporal feature is only covariate with a large contribution to the temporal variance (See Figure S2). We observe that the means of features change throughout the study for their respective classes (See Figure S3). However, covariate shift is *not* the main source of dataset drift in the wearable features. Instead we attribute it to the fact that the likelihood of acquiring COVID-19 shifts as prevalence of COVID-19 changes, i.e. there is a prior probability shift [40]. We address this in model development by selecting thresholds on a prospective validation set, instead of using a randomly selected validation set. We found this to be essential to attain reasonable performance on future data during the ebbing prevalence of COVID-19 with a single season of data (See Figure S8).

### COVID-19 detection using only wearable data

For any prospective week, a model is trained to differentiate symptomatic COVID-19 days from symptomatic ILI days, or healthy days prior to illness. When models are evaluated on prospective data, we find that the margins of separation for COVID-19 vs. non-COVID-19 ILI & healthy classes is small. These weekly results are reported in Figure 4a for XGBoost. By nature of the `FLUCOVID` study design, only individuals who eventually experience an ILI are included. For any given week using wearable devices to directly detect COVID-19 achieves a sensitivity of 0.52 (0-1, 95% CI), with a false positive rate of 0.49 (0.04-0.99, 95% CI). The test set sensitivity will not change as more negative controls are added to the model, though all other metrics (specificity, positive predictive value, negative predictive value, AUROC, and AUPR) are affected by the negative class proportion. The reported scores are conservative, as we have a disjoint set of participants *and* non-intersecting date ranges between the training, validation, and test sets. In reality only a small proportion of participants would be expected to enter or leave the survey each week, meaning the deployment performance would only be harmed by lack of generalisation through time, rather than lack of generalisation through time and across participants.

**Figure 4.**
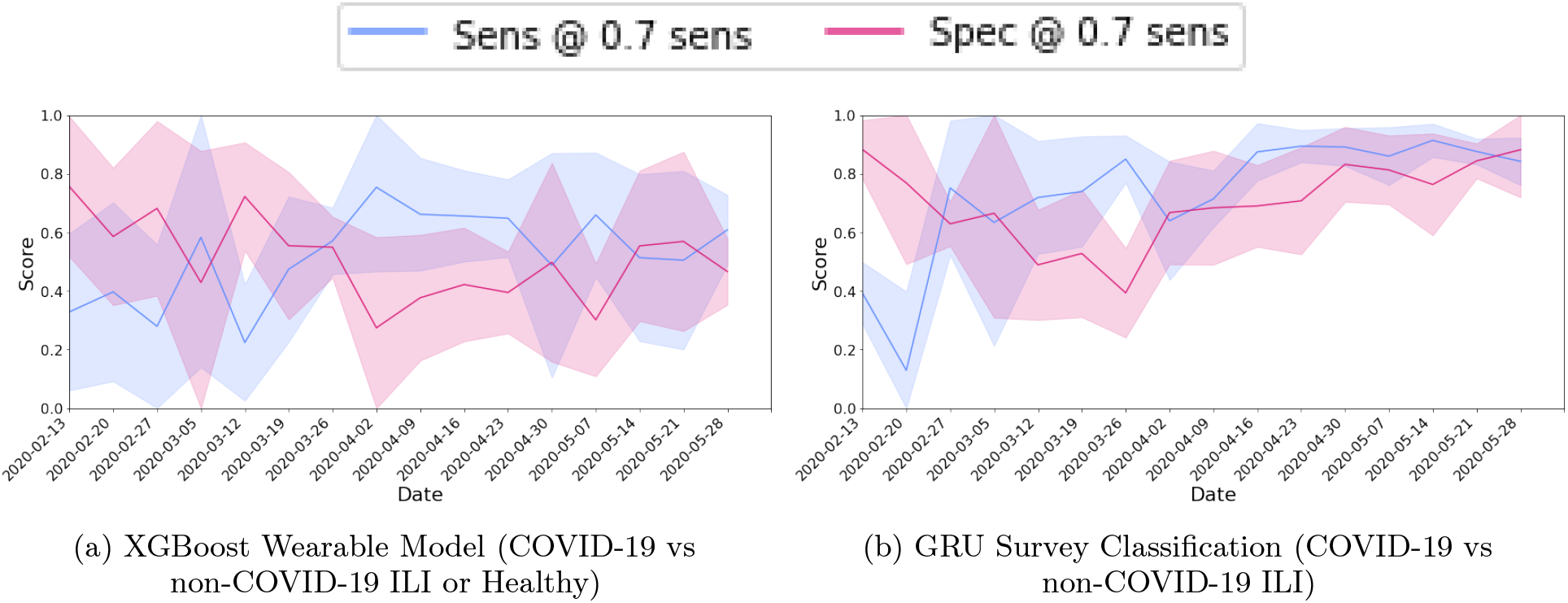
Wearable vs. survey model performance: Performance of two data modalities, wearable data or survey data, for directly predicting COVID-19 each week. Results are shown as mean ± standard deviation

### COVID-19 detection using only survey data

Ultimately, it could be easier to differentiate between COVID-19, ILI, and healthy days using daily surveys [50]. However, long-term engagement in digital health studies is a burden to the model’s beneficiaries [47]. Survey models currently upper bound mobile COVID-19 diagnostics. Though, surveys will still fail to detect pre-symptomatic, asymptomatic, and possibly mild symptomatic COVID-19 detection. Recall that for COVID-19, 17% of cases are estimated to be asymptomatic [5; 39], which is comparable to the rate of asymptomatic influenza [32]. Both asymptomatic COVID-19 [30] and asymptomatic influenza [21] shed viral RNA. For those who eventually develop symptoms, the relatively long incubation time of SARS-COV-2 is problematic as 44% of transmission occurs during pre-symptomatic phase [17], though active viral load peaks around symptom onset [9]. These sources of transmission are unable to be prevented with symptom-based screening alone.

Our data is collected such that a participant would not be asked to complete a survey if they did not have symptoms. The task for this survey model is to detect which symptoms are a result of COVID-19, and which can be presumed to be non-COVID-19 ILI. Models trained on the daily symptom history and demographic covariates to distinguish COVID-19 positive individuals from other ILI. The top performing model, a GRU, achieves this task with an AUROC of 0.78 ± 0.19 and an AUPR of 0.28 ± 0.11 for a prospective week, on a disjoint set of participants. The sensitivity and specificity of this method is shown in Figure 4b. In comparison, the linear model on a curated set of features as reported in [34; 50] yielded AUROC of 0.62 and AUPR 0.03 on our entire `FLUCOVID` survey data. Since the nature of the task is to determine if symptoms are caused by COVID-19 or not, and since all ILI cases are included in the study by design, the reported metrics would remain valid even if additional healthy individuals were included.

### Triggering survey model using wearable predictions for COVID-19 detection

For any sensitivity chosen on the wearable model, we always improve the likelihood that the individual has COVID-19 (See Figure S10). Hence for a permissive sensitivity, we can dismiss many healthy individuals, which minimises reliance on continued survey engagement. The performance of the XGBoost and GRU-D wearable models, paired with a GRU survey model are shown in Figures 5a & 5b. For an average week, the performance on a disjoint set of participants can be expected to be 0.50 (0-0.74, 95% CI) sensitivity and 0.79 (0.53-0.98, 95% CI) specificity for the XGBoost architecture on the wearable model combined with the GRU survey model.

**Figure 5.**
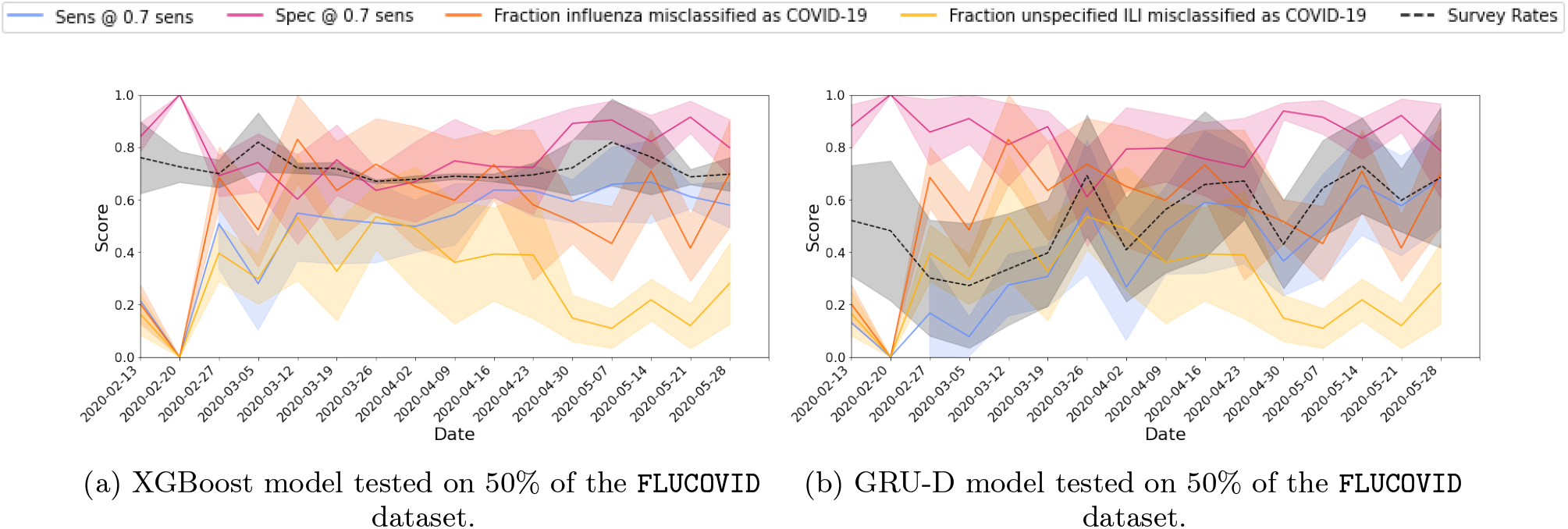
Combined wearable and survey model performance: Performance of the Covid-19 Wearable+Survey detection models in the `FLUCOVID` dataset (top) and `COVID2020` dataset (bottom) for both XGBoost (left) and GRU-D (right) wearable models.

However, given a positive prediction at any date near or during the onset of infection, participants could have been directed to get further confirmatory testing. Any positive prediction leading up to symptom onset could elicit behavioural change such as staying home from work or school, or getting a PCR test. Under this assumption, false negative results that follow positive predictions do not have such a profound impact. We consider a cumulative score automatically classifies a positive score any of the past 7 days has been classified as positive. We hypothesise that this would increase the sensitivity (by decreasing the number of FN) while decreasing the specificity. This nuance in reporting leads to an expected weekly sensitivity of 0.65 (0.19-0.87, 95% CI) and specificity of 0.69 (0.41 -0.97, 95% CI) on prospective test sets on `FLUCOVID` data. Roughly 63.5% of COVID-19 cases are detected at symptom onset (47.7% for non-COVID-19 ILI), and 68.9% of cases are captured by day 3 of symptoms (55.8% for non-COVID-19 ILI), before many individuals would be compelled to take a PCR test and receive results (See Figure S11).

The wearable component for this prediction scheme prompts surveys 65% of the time (a 35% reduction in survey burden as compared to daily symptom reporting). The sensitivity of the wearable-only model (any ILI detection) is 0.71 (0.63-0.88, 95% CI), and a specificity is 0.35 (0.15-0.46, 95% CI). Note that thresholds are chosen on a prospective validation set where the sensitivity is 0.7. The performance on the wearable portion of the combined model is considerably better than the wearable models for direct COVID-19 prediction that were distinguishing COVID-19 from ILI since the combined approach does not have to differentiate COVID-19 from ILI, and the ILI labels are more frequent in our data set.

### Minority groups are not disparately impacted by wearable plus survey COVID-19 detection

The pandemic has a disparate impact on minorities. One meta-analysis found Black and Asian groups were more likely to contract COVID-19 than their White counterparts [58]. These results are in agreement with several other studies for Black [23; 42; 49], Hispanic, Asian, and North American Indigenous minorities [22]. We look at the performance of the combined wearable and survey model subset by race and gender, where subgroups have more than 1000 members. These results are shown in Table I. Results show that the differences between male and female subpopulations are significant for both specificity and sensitivity, but there is a tradeoff: for the same decision threshold, one group experiences a higher sensitivity and lower specificity, whereas the other group experiences the opposite effect. This pattern is also observed in the black participants where the model sensitivity on black participants is 12% less than the overall sensitivity, but the specificity is 10% higher with the same threshold. In models where the `week-of-year` is omitted, the sensitivity difference is abridged. Our results also show that the model’s sensitivity is significantly higher on the Asian subpopulation compared to the non-Asian population. The sheer number of non-COVID-19 days renders all of the examined subpopulation specificities significant when compared to the rest of the population. The Asian, Black, and Hispanic minority subpopulations have specificities exceeding the population mean, whereas the white subpopulation specificity is below that of other groups. Overall, the model performance does not exacerbate existing inequalities of the COVID-19 pandemic noted in previous studies [23; 42; 49; 58]. However, in specific use contexts it may be essential to leverage post-processing techniques to achieve demographic parity or equalized odds.

## DISCUSSION

### Importance of including imperfect data

All of the 32, 198 participants in the `FLUCOVID` study had at least one ILI event and consented their wearable survey data. Participants have varying degrees of device wear time. Some may only take it off to charge it (nearing 100% wear time when acquiring signals for an entire day) whereas others have sporadic usage. Previous studies require a minimum device wear time for their cohort selection [35; 43], and all participants who do not meet this requirement are dropped. We instead choose models that can implicitly handle missingness. When we gradually eliminate patient-days based on their percent of missingness, we find that separation between ILI and healthy classes diminishes (See Figure S12). Intuitively, this suggests that including missingness makes the task more difficult. By the nature of the task, abstaining from making a prediction is synonymous with classifying a participant as healthy. In order to compare wearable device model performance to other diagnostic measures, such as PCR tests, missing data cases cannot be neglected in test data. We replace these missing predictions with a classification of healthy when evaluating models on test data.

It is important to note that all labels in the dataset have been created using self-reported symptoms, onset dates and recovery dates. When the time between the self-reported date of disease recovery and the survey response is more than one week, model area under the precision recall curve (AUPR) drops considerably corresponding to the reduction in positive cases, however the sensitivity and specificity are not overly perturbed by delays in reporting (See Figure S7). The deteriorating AUPR is driven by a decreasing positivity rate.

**Table I.**
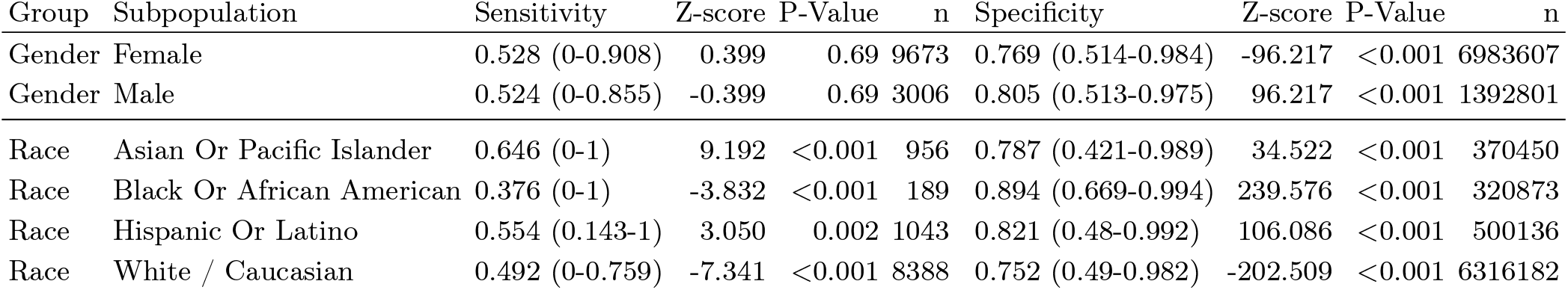
Combined wearable and survey model performance for COVID-19 detection by race or by gender for cohorts with more than 1000 participants. Results are reported as score (95% confidence interval). Z-score calculated as difference in binomial proportions (Wald test) to other groups (e.g. Asian vs non-Asian). This includes `week-of-year` and null survey answers are assumed to be healthy predictions.

### Reporting performance across the entire study

Our reporting differs from existing literature in that we report across the entire timeline whereas other approaches [35; 38; 43; 50] report classification results for the windows around the onset. A machine learning model trained to detect ILI would be expected to have a detectable change in outputs corresponding to an increase in detection around the time of ILI symptom onset. It is tempting to validate the machine learning model by aligning participants in the test set by the date of onset to confirm this. Previous research has demonstrated that model performance will be artificially inflated unless an outcome-independent reference point is used [54]. Prior wearable work calculates AUROC and the ROC is plotted after having discarded a week of data before the reported date of onset, and excludes dates more than 7 days after onset and earlier than 21 days prior to onset [43; 50]. However, even a small false positive rate can completely neutralise the perceived utility of the model for an imbalanced task such as determining ILI days vs healthy days. For posterity, we also aggregate our results in this fashion in Figure 6b. A full comparison of our model performance to published models, under their own evaluation strategies can be found in Table S7, though we do not encourage communicating model performance according to these metrics as they do not reflect model utilisation.

**Figure 6.**
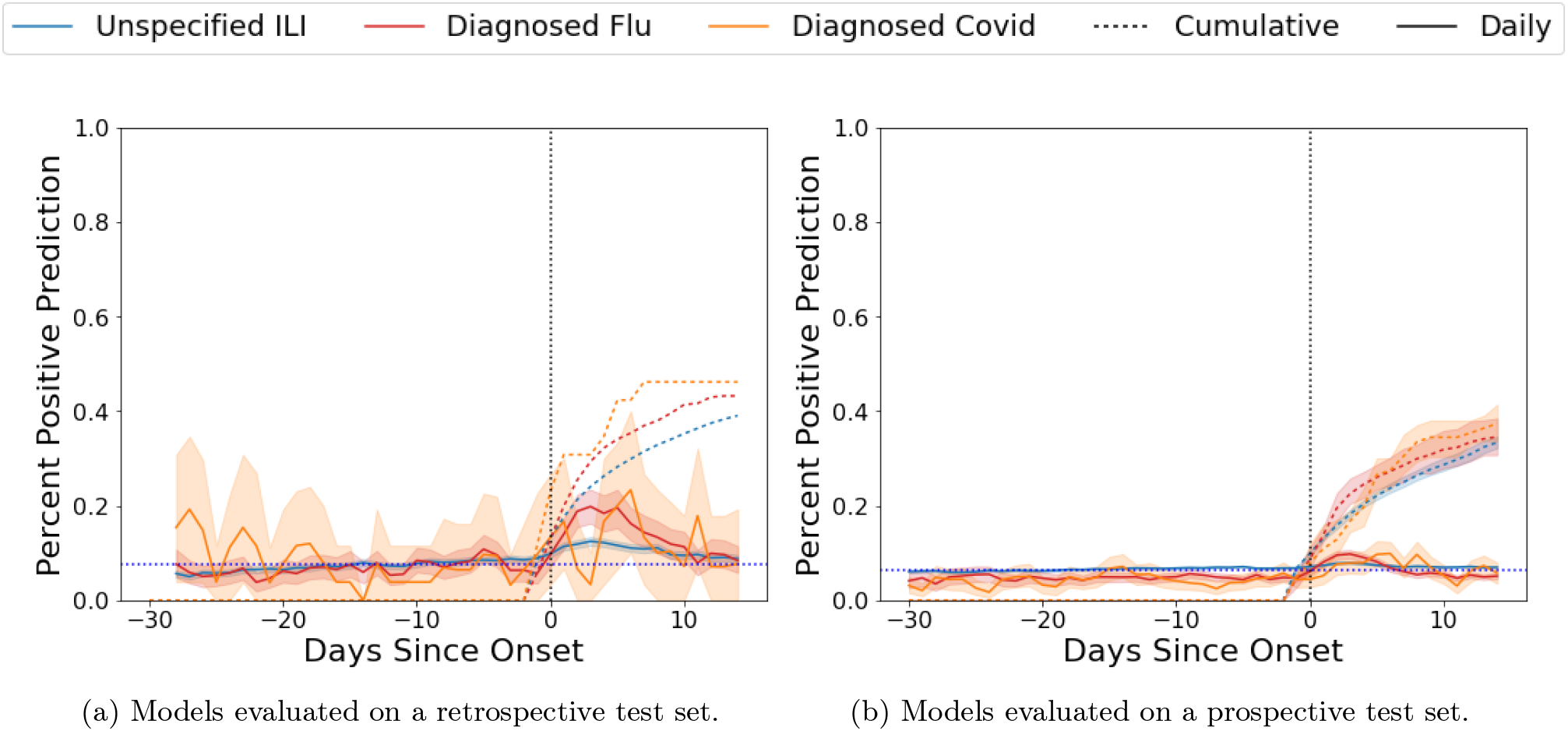
Positive calls around symptom onset: XGBoost model performance for detecting any ILI or COVID-19 both retrospectively and prospectively and grouped relative to the symptom start date (day 0). Note that elevated percent predicted after day 0 indicates better performance under our labelling scheme.

A similar design was followed where nighttime respiratory rate, RHR, and heart rate variability (HRV) were used to train a model to classify the onset of COVID-19 [35]. To accommodate the class imbalance between sick and healthy days in the training set, sick days were synthetically oversampled by adding noise. The model captured 80% of the COVID-19 positive individuals, but the same model detected 34% of individuals who exhibited symptoms but tested negative for COVID-19. This is understandable as the model was not specifically trained to discern between COVID-19 positive and negative cases. However, this evaluation should be observed cautiously. The number of sick days, and the number of healthy days in the objective are arbitrarily chosen; only days within -30 to -14 range were used in the validation sets as negative labels (For COVID-19, 99% of symptomatic cases will show symptoms within 14 days after exposure [2; 29]). For those that eventually got COVID-19 the FPR was 4.7% and those who eventually had ILI had a FPR of 5%. The FPR of 4.7% corresponds to a COVID-19 prediction once every 21 days, or 17 false positive calls per year for a healthy individual [35].

In another study, an online outlier detection method was shown to correlate with a post-hoc/offline outlier detection method on heart rate, heart rate to steps ratio, and sleep features to detect the onset of symptoms for 32 SARS-COV-2 positive individuals [38]. Specifically, their method correctly identified an outlier region within 14 days of SARS-COV-2 symptom onset for the majority of users. When we applied this outlier detection method to a subset of our `FLUCOVID` cohort, we were also able to find a detection event within 14 days of symptom onset for most individuals. However, the sensitivity of this approach was strongly influenced by the amount of data that was recorded before and after the SARS-COV-2 symptom onset date. For example, when we (artificially) decreased the amount of data by 14 day increments in our study, the symmetric sensitivity (a detection event within 14 days before/after onset) rose from 63% to 100%, and the one-sided sensitivity (a detection event within 14 days before onset) rose from 40% to 61%. Reducing the amount of recorded data around onset increased the positive predictive rate from 27% to 30%, as well as lowered the p-value calculated relative to a null detection proportion from 3% to <1%. Constraining the detection window size over which any ML method is evaluated will artificially increase sensitivity, specificity, and statistical significance. Outlier detection results are unlikely to generalize for small time frames. For COVID-19 the median time from exposure to onset is 4-6 days [2; 29; 65], and 97.5% of symptomatic cases occur within 11.5 days after exposure [29]. If there is exposure earlier than 14 days to prior to symptom onset, it is unlikely to be a true positive. Influenza has an even shorter median incubation time, with only 1.4 or 0.6 days until influenza A and influenza B symptom onset, respectively [31].

Grouping the predictions by proximity to ILI cases can hide real trends in the data. Calculating the AUROC around onset is highly influenced by the class balance [46] which in this setting is contrived. Prior commentary is available on the attribution of important baselines, such as prevalence [13]. The likelihood of contracting COVID-19 is non-stationary because the transmissability of COVID-19 changes in response to public health measures, the number of infected cases and the circulating strains [1]. Evaluation procedures must be agnostic to these effects. This type of dataset shift is known as prior probability shift [40], which warrants careful calibration of machine learning models or additional training techniques [52]. In this study we have focused on evaluating our models and communicating model performance that is reflective of deployment under these circumstances.

### Account for ILI cases when building a COVID-19 detector

It is essential that studies account for ILI cases when building COVID-19 wearable detectors [53]. When applying machine learning models to aid in the detection of emergent diseases we must use training data which are representative of data in a deployment scenario. An individual with influenza may share several characteristics of COVID-19, despite never being exposed to the SARS-COV-2 virus. There is an overlap between COVID-19 and influenza in both symptoms and autonomic responses which cannot be ignored when reporting the specificity of COVID-19 diagnostic tests. Fortunately, these overlapping symptoms present with different frequencies. For example, COVID-19 patients are significantly more likely to have ageusia (loss of taste), fever, anosmia (loss of smell), myalgia/arthralgia (aching body), diarrhea, and nausea [7; 34; 63]. The symptoms which are more prevalent in COVID-19 negative ILI patients are cough and sore throat, though only sore-throat is statistically more likely to be associated with influenza [63]. The associations of these symptoms with COVID-19 have been corroborated in other studies [10; 43]. Symptom based COVID-19 detection is somewhat able to distinguish COVID-19 positive patients from other respiratory illnesses using a simple logistic regression model [6]. However, when determining the chance of a positive PCR COVID-19 test given that a patient presents symptoms, the major drivers of the prediction were travel history and case proximity in an early stage of an outbreak (not fever or cough) [10]. These results will vary based on the richness of the feature set, the accuracy of symptom recording, the size of the cohort, and the progression of the pandemic in the region. Using wearable device data in place of symptom based screening exchanges improved frequency of monitoring with further abstraction from the non specific symptom features.

### COVID-19, mild symptomatic COVID-19, and ILI differentiation

With COVID-19, mild symptomatic COVID-19, and ILI all concurrently present, it is essential that models can differentiate between them. Differentiating between COVID-19 days and healthy days tends to be the easiest task amongst the subclasses of COVID-19, influenza, and unspecified ILI, alongside the healthy class (Figure S9). It is considerably more difficult to differentiate between unspecified ILI and healthy cases.

Unspecified ILI cases in the `FLUCOVID` surveys subside at a proportionately slower rate than the CDC’s reported values [8] (See Figure 1). We suspect that these ILI cases may be undiagnosed, or misdiagnosed “mild symptomatic COVID-19”. Amongst the 8743 participants who reported ILI symptoms and *did not* test positive for SARS-COV-2 between April 1, 2020-June 1, 2020, 86% received a negative COVID-19 test within -2 and 21 days after symptom onset (14 days post symptoms +7 days reporting delay). Only 6% of patients who didn’t have COVID-19 tested positive for influenza in the `FLUCOVID` dataset. Several of these included participants who had not been tested for COVID-19. Less than 1% of participants in this time period experienced symptoms, but did not get tested for influenza or COVID-19, have had not previously tested positive for COVID-19, yet reported having close contact with a COVID-19 case within the 14 days prior to symptom onset. This small proportion of untested, COVID-19 contact, unspecified ILI cases equates to roughly 10% of the confirmed COVID-19 cases during the same period. If any of these untested cases are actually COVID-19 it could decrease the specificity in our reported results, as the TPs would be disguised as FPs. This coincides evidence of individuals in the USA who had been infected with SARS-COV-2 based off of the presence of antibodies, yet had not been reported [3] (though these results must be interpreted cautiously [56]). These individuals would still be capable of transmitting virus, however if we followed the convention of previous works, unspecified ILI patients would have been excluded from the dataset. The signal that would be learned under that convention would be the joint probability of getting a PCR test *and* that test being positive for SARS-COV-2 [15]. We want to discern the latter unconditioned on getting a PCR test.

### Limitations

There are several limitations of this study which would affect deployment-level performance. Most importantly, our training dataset does not contain participants who are healthy throughout the entire duration of the study. Upon deployment, the PPV, NPV, specificity, AUROC, and AUPR will all be affected since these metrics involve the calls made on COVID-19 negative participants. In addition, the participants consenting in these studies may not represent the population which this would be deployed for. Both cohorts are comprised of primarily female, and primarily white demographics. This could be attributed to the selection bias in those who wish to participate, or the selection bias of individuals who own a wearable device. We find that symptom-based screening substantially outperforms wearable device screening in differentiating between COVID-19 cases and non-COVID-19 ILI cases. The limitation of symptom screening is that it negates the ability of the wearable devices to identify positive COVID-19 cases prior to symptom onset. Both models could be improved from enriched features. For example changes in heart rate variability metrics have been shown to be associated with COVID-19 [19; 35; 43]. Our features are summarised on the day level. More granular time intervals could improve identification of ILI signatures.

Improvements could be made to the modelling such as using side information from the survey data to improve the representational capacity of the wearable-only model, or a one-time demographic survey could be required so that wearable devices can leverage participant attributes to improve performance. A further reduction on the reliance of surveys could be achieved through objectives that learn to defer to surveys when the wearable-model is uncertain [41]. Specific modelling techniques could be applied to address dataset shift due to frequently changing lockdown status or changing prevalence [52]. The prospective results on this study report performance on a disjoint test set, which is likely to overestimate the performance gap caused by generalisation across time by encompassing the generalisation across participants. Users could potentially benefit from having personalised models to capture outlying personal patterns [64], though global patterns would need to be leveraged for COVID-19 detection since it does not occur more than once per individual in our datasets. Finally, changes in how these predictions are used could impact their performance. Perhaps tests or treatments are scarce and only allocated to the top-*k* individuals. In another use case, not providing a prediction when there is insufficient data would improve the performance of the model. This could be useful in a context where a negative prediction is required to attend class in person (whereas positive and null predictions would dictate remote attendance).

## ONLINE METHODS

We collected Fitbit activity data, demographic baselines, and weekly symptom surveys from 33777 participants from November of 2019 through August, 2020. All users reside in the United States. We refer to this dataset as `FLUCOVID`. The characteristics of this dataset are described in greater detail in the following sections. For details on the collection of data, we refer the reader to our previous work [53].

### Demographic Data

Participants consented data for the study which includes age, education, ethnicity, children and relationship status, height, weight, and location. In addition to these demographic variables, comorbidities are available including asthma and COPD (amongst others). Demographic data and comorbidities are not used in training wearable models, because they are not available in the wearable device data. This allows us to cast the widest net in COVID-19 detection (sensitivity) at the expense of specificity. The characteristics of the data can be found in Table II.

**Table II.** `FLUCOVID`: Demographics of the Large-scale Flu Surveillance study for survey and wearable cohorts

|  | Survey cohort |  | Wearable cohort |  |
| --- | --- | --- | --- | --- |
|  | Total | COVID-19 | Total | COVID-19 |
| **Gender** |  |  |  |  |
| Female | 77.1% | 68.9% | 82.2% | 77.7% |
| Male | 21.7% | 29.4% | 16.5% | 20.6% |
| Other | 0.6% | 0.2% | 0.6% | 0.3% |
| **Race and Ethnicity** |  |  |  |  |
| American Indian Or Alaska Native | 0.5% | 1.5% | 0.5% | 2.3% |
| Asian Or Pacific Islander | 5.5% | 8.9% | 4.4% | 6.5% |
| Black Or African American | 4.3% | 6.0% | 3.8% | 5.4% |
| Hispanic Or Latino | 7.2% | 10.1% | 5.9% | 10.7% |
| Unavailable | 1.8% | 2.7% | 1.7% | 2.8% |
| White / Caucasian | 72.2% | 57.4% | 74.3% | 60.7% |
| **Sample size** | 81088 | 823 | 33777 | 354 |

### Wearable Device Data

Our wearable features contain 48 covariates derived from heart rate monitoring, step tracking, and sleep tracking features for each day (Table S1). We compare several data preparations, including regularly vs irregularly indexed data [20], population normalisation vs. individual normalisation [51], and forward filling imputation vs. no imputation. A complete description of our data preparation is available in Supplementary A. When the effect is insignificant, or the effect size is small between preparation tasks, we err on the side of including as many patient-days as possible. Unless otherwise specified, we use regular indexing, population-level normalisation, and forward-filling imputation while reporting our results.

### FLUCOVID Survey Data

Each participant in the `FLUCOVID` cohort experienced at least one influenza-like illness (ILI) event between November 1, 2019, and June 1, 2020. Of the 32198 participants with *both* surveys and wearable data in this cohort, 2554 had medically diagnosed influenza, 204 had COVID-19, and the remainder had unspecified ILI. 27% of users reported more than one ILI event in the study. All of our labels in the survey are derived from self-reported symptoms and test results. Each week, participants are asked to recall their symptoms in the past 7 days. They will note which days they experienced flu related symptoms, COVID-19 related symptoms, and complications with chronic diseases. ILI onset and recovery dates are also reported by the participant. In addition to illness, there are questions pertaining to self-isolation, air travel, and household members with ILI or COVID.

### Model Training

We opt for machine learning models that can implicitly handle missingness, namely, we use GRU-D [11] or XGBoost [12] models. Models are exclusively trained on the `FLUCOVID` dataset such that 35% of participants fall into the training set, 7.5% in the validation set, 7.5% in the held out random retrospective test set, and 50% in the held-out prospective test set (to preserve prevalence). Features are collected on a daily basis, and predictions are made at the same frequency. A complete description of the features, and their preprocessing can be found in Supplementary A. For the XGBoost model, we use the past 7 days of data (inclusive of the prediction day), flattened into a matrix 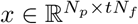, where *N*_*p*_ is the number of participants, *t* is the number of days, and *N*_*f*_ is the number of features. This flattening procedure is rolled across every time-point for each participant. Hyper-parameter selection was performed to minimise the validation set loss whenever a new model is trained. The GRU-D model ingests data in the format of 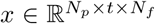 as it is recurrent. The GRU-D model used the validation set for early stopping. Because of this, Survey cohort Wearable cohort Total COVID-19 Total COVID-19 different weights were learned for the model which is stopped on the concurrent portion of the validation set for randomised testing as opposed to the prospective portion of the validation set for prospective testing. The comparisons between randomised and prospective evaluations for the XGBoost model use the *same* weights in our results whereas the same comparisons for GRU-D do not. The participant splits between XGBoost and GRU-D models are shared, so the performance can be directly compared between the two models.

The survey models used a subset of features that are common for COVID-19 screening questionnaires as seen in Table S2. Notably, several of these features are significantly different between those who tested negative for COVID-19, and those who tested positive. Even still the relative abundances of behaviour and travel are similar. Statistical significance does not promise separability during inference. These features, combined with demographic variables are ingested into a GRU model. Due to the nature of the surveys, days where no answer is provided are assumed to be symptom free. Models are trained only where symptoms are reported, to differentiate between COVID-19 and non-COVID-19 ILI using all symptoms and behaviour patterns observed to date.

Choosing thresholds on all historical validation data does not generalise to the risks that are predicted in the following prospective week. Due to the class imbalance, outcome probabilities (logits) are poorly calibrated. A prospective validation set is necessary to select thresholds which will be calibrated in the next week. Through this process we found the choice of threshold for a given sensitivity on that validation set would better estimate the sensitivity we would observe on the subsequent test set (See Figure S8b). In our case this can be attributed to inconsistency of the target distributions (prior probability shift) [40] seen in Figure 1. For randomised test sets, thresholds were drawn on a validation set that was concurrent with the training and test sets.

## Data Availability

The completed coded curated study data can be requested by qualified researchers via the Sage Synapse platform: https://www.synapse.org/#!Synapse:syn22891469/.

## ACKNOWLEDGEMENTS

Funded by NIH, The National Cancer Institute (NCI) and the National Institute of Biomedical Imaging and Bioengineering (NIBIB) AWARD N. 75N91020C00034.

## DISCLOSURE OF POTENTIAL COMPETING INTERESTS

Luca Foschini is co-founder of Evidation Health, a company that powers research studies with PGHD (Person-Generated Health Data).

## AUTHOR CONTRIBUTIONS

BN, JH, and RK completed modelling, experimental design, visualisation, and reporting. ED completed statistical tests and re-implemented a prior work. RK, JI, and AS designed data collection and prepared the datasets. SN provided input on COVID-19 related literature and survey feature selection. MG, LF, and AG regularly advised on the project and determined outcomes.

---

**Figure S1.**
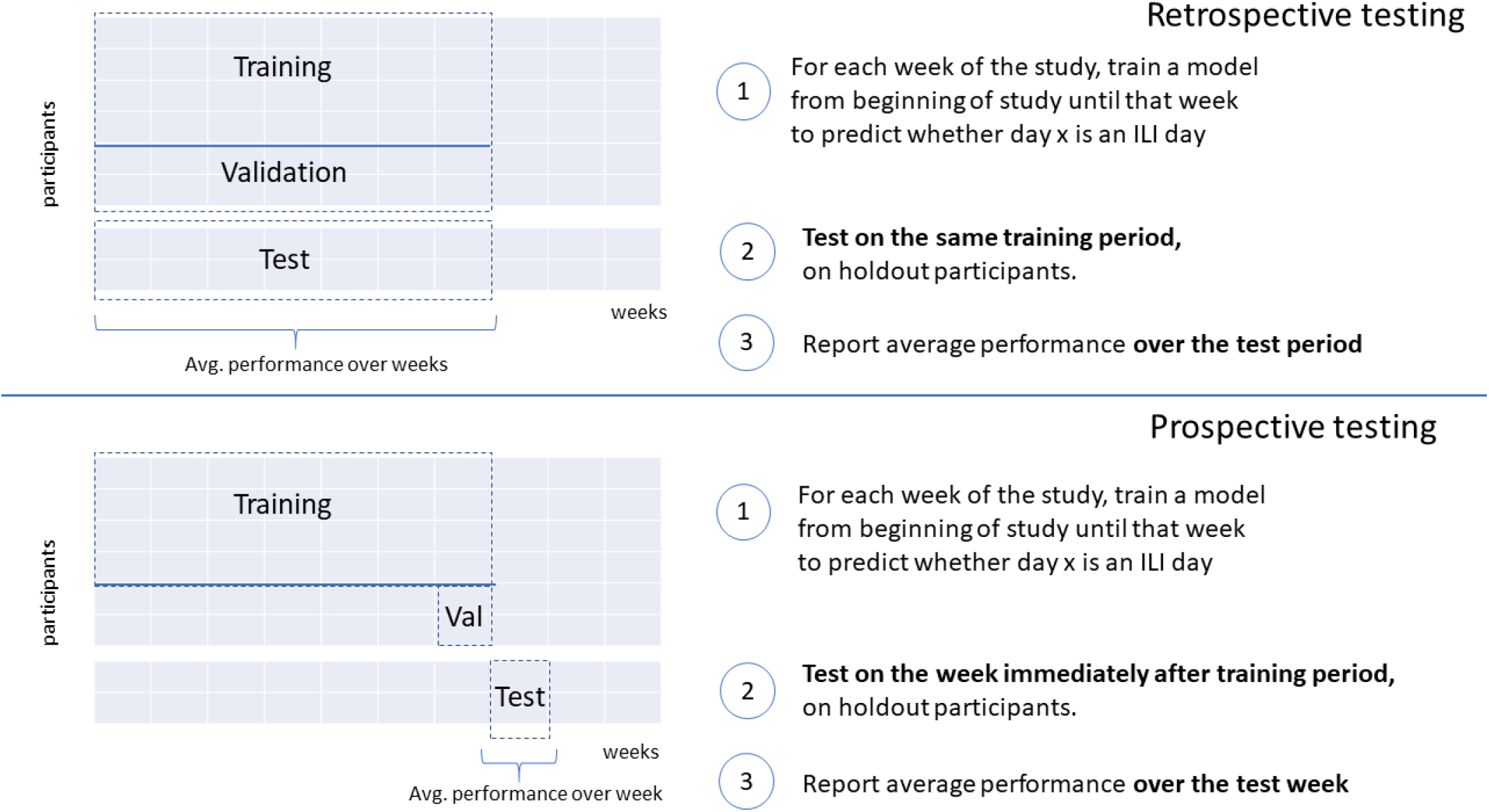
Retrospective vs. Prospective testing setup

## Appendix A: Wearable Features

The day-level features available to the wearable model are listed in Table S1.

We used t-SNE and PCA to investigate the feature dependence on time. We show the results in Figure S2.

## Appendix B: Survey Features for FLUCOVID

We show the survey features that are available to our survey model for the participants in the `FLUCOVID` dataset in Table S2.

## Appendix C: Data Preparation

We compared several data preprocessing steps: irregular vs. regular sampling [20], population-level vs. individual-level normalisation [51], and forward-filled imputation vs. no imputation. When a day has no observed data, the data is re-indexed to obtain a regular interval (Table S4), or left alone to retain an irregular interval [20] (Table S3). Examples for irregular sampling and regular sampling are shown in Tables S3 & S4, respectively. We found that the sampling interval did not substantially affect retrospective model performance on randomised training. validation, and test splits (See Figure S4).

Next, the data normalisation is considered. For the z-score normalisation the rolling average and standard deviation is calculated for each of the patients, and each of the features, across 28 time-points of patient data. For the regularly sampled data, this corresponds to 28 days of data (some of the data will be missing), whereas for the irregularly sampled data, it simply means the previous 28 days in which any measurement had occurred (all of the data will have been observed). We use the mean and standard deviation from the 28 day period to calculate the individual’s z-score (only where at least 14 observations are made) similar to the procedure followed in previous flu monitoring pipelines [51]. Our intention is to avoid making predictions based on an individual’s deviation from the population average [13], and instead make predictions based on an individuals own internal consistency. The mean and standard deviation of the individuals data is buffered by a 7 day gap. This means that on the first day of ILI symptoms, the z score is unencumbered by the most recent 7 days of data to avoid the bias from the viral incubation period. however, illnesses lasting over a week will start to normalise the elevated resting heart rate values. A larger gap, or a longer data requirement period could be administered at the expense of requiring a longer participation period before a prediction can be made. This calculation is shown in equation C1.

**Table S1.** Features Available from Wearable Device

| Feature Name | Type | Unit | Source | Description |
| --- | --- | --- | --- | --- |
| heart rate bpm | heart rate | bpm | FBDL | daily resting heart rate |
| sleep seconds | sleep | seconds | FBDL | daily total sleep |
| walk steps | steps | count | FBDL | daily total steps |
| active Fitbit awake sum | wear time | minutes | FBML | daily sum of awake wear-time |
| active Fitbit sum | wear time | minutes | FBML | daily sum of wear-time |
| heart rate asleep max | heart rate | bpm | FBML | max asleep daily heart rate |
| heart rate asleep mean | heart rate | bpm | FBML | mean asleep daily heart rate |
| heart rate asleep stddev | heart rate | bpm | FBML | stddev asleep daily heart rate |
| heart rate awake max | heart rate | bpm | FBML | max awake daily heart rate |
| heart rate awake mean | heart rate | bpm | FBML | mean awake daily heart rate |
| heart rate awake stddev | heart rate | bpm | FBML | stddev awake daily heart rate |
| heart rate mean | heart rate | bpm | FBML | mean daily heart rate |
| heart rate not moving max | heart rate | bpm | FBML | max non-moving daily heart rate |
| heart rate not moving mean | heart rate | bpm | FBML | mean non-moving daily heart rate |
| heart rate not moving stddev | heart rate | bpm | FBML | stddev non-moving daily heart rate |
| heart rate perc 25th | heart rate | bpm | FBML | 25th percentile daily heart rate |
| heart rate perc 50th | heart rate | bpm | FBML | 50th percentile daily heart rate |
| heart rate perc 5th | heart rate | bpm | FBML | daily resting heart rate |
| heart rate perc 75th | heart rate | bpm | FBML | 75th percentile daily heart rate |
| heart rate perc 95th | heart rate | bpm | FBML | 95th percentile daily heart rate |
| heart rate resting heart rate | heart rate | bpm | FBML | daily resting heart rate |
| heart rate stddev | heart rate | bpm | FBML | stddev daily heart rate |
| sleep main start time | sleep | hr | FBML | time of day of sleep start |
| sleep asleep mean | sleep | fraction | FBML | fraction of sleep spent asleep |
| sleep asleep sum | sleep | minutes | FBML | minutes in main sleep spent asleep |
| sleep awake mean | sleep | fraction | FBML | fraction of sleep spent restless-awake |
| sleep awake sum | sleep | minutes | FBML | minutes in main sleep spent restless-awake |
| sleep awake regions count | sleep | count | FBML | number of distinct contiguous regions of restless-awake sleep state |
| sleep main efficiency | sleep | fraction | FBML | Fitbit sleep efficiency score 0 to 1 |
| sleep nap count | sleep | count | FBML | number of naps in a day |
| sleep really awake mean | sleep | fraction | FBML | fraction of sleep spent really-awake |
| sleep really awake sum | sleep | minutes | FBML | minutes in main sleep spent really awake |
| sleep really awake regions count | sleep | count | FBML | number of distinct contiguous regions of really-awake sleep state |
| sleep sleeping sum | sleep | minutes | FBML | minutes in Fitbit sleeping staus |
| sleep total asleep minutes | sleep | minutes | FBML | total asleep minutes for day |
| sleep total in bed minutes | sleep | minutes | FBML | total minutes in bed for day |
| steps count | steps | count | FBML | number of minutes of day with steps data |
| steps dec time max | steps | hr | FBML | time of day of last daily steps |
| steps dec time min | steps | hr | FBML | time of day of first daily steps |
| steps dec time max rolling 6 first | steps | hr | FBML | time of day when first max rolling-6-min steps taken |
| steps light activity sum | steps | count | FBML | total light-activity daily steps |
| steps mvpa sum | steps | count | FBML | total moderate-vigorous physical activity daily steps |
| steps not moving sum | steps | minutes | FBML | "total daily non-moving minutes & where steps-streak <= 10" |
| steps rolling 6 sum max | steps | count | FBML | max rolling-6-min daily steps |
| steps sedentary sum | steps | count | FBML | total light-activity daily steps |
| steps streaks countDistinct | steps | count | FBML | number of distinct step-minute streaks in a day |
| steps sum | steps | count | FBML | total daily steps |

**Figure S2.**
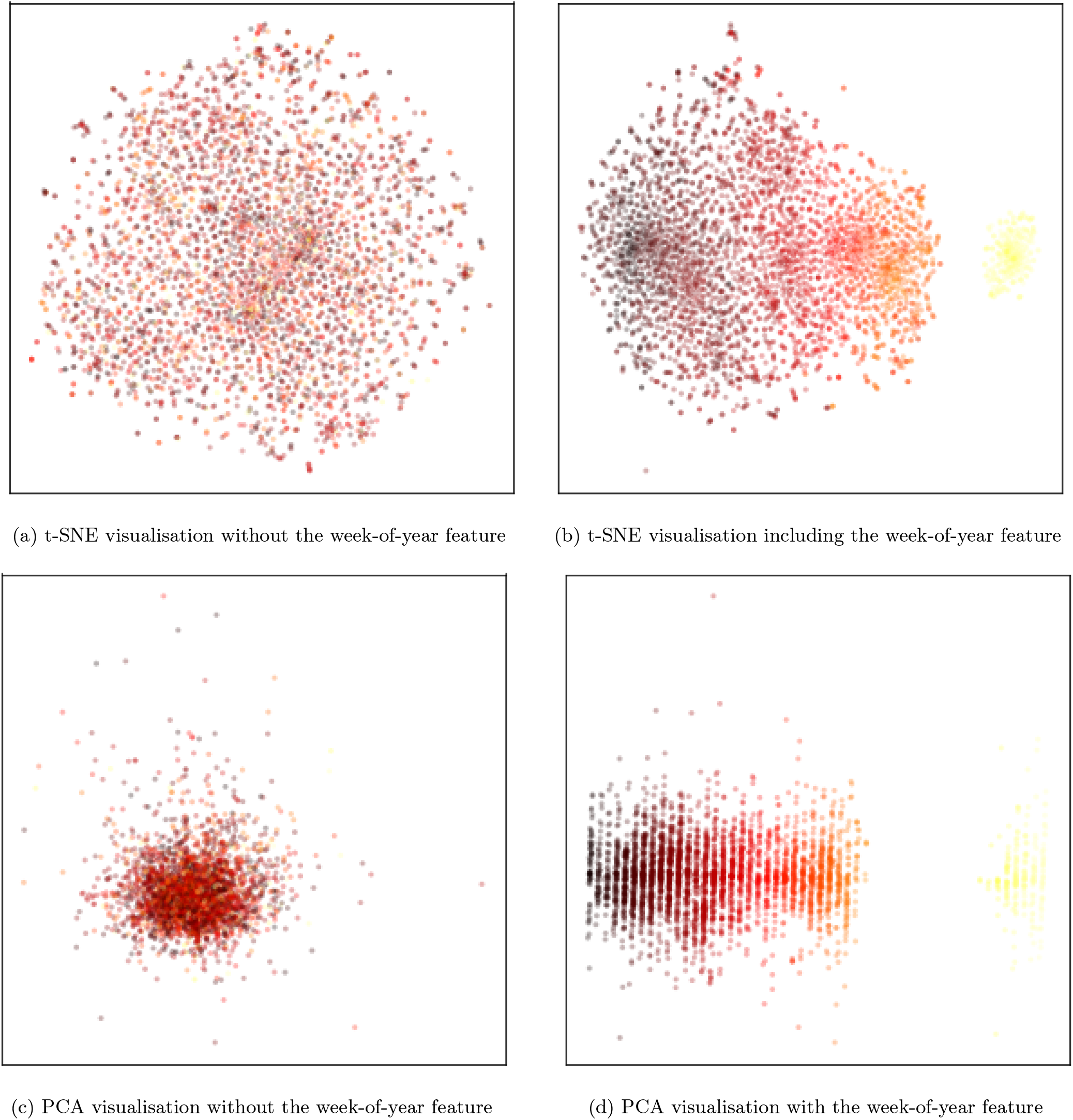
Visualisations of wearable features coloured by `week-of-year`.

**Figure S3.**
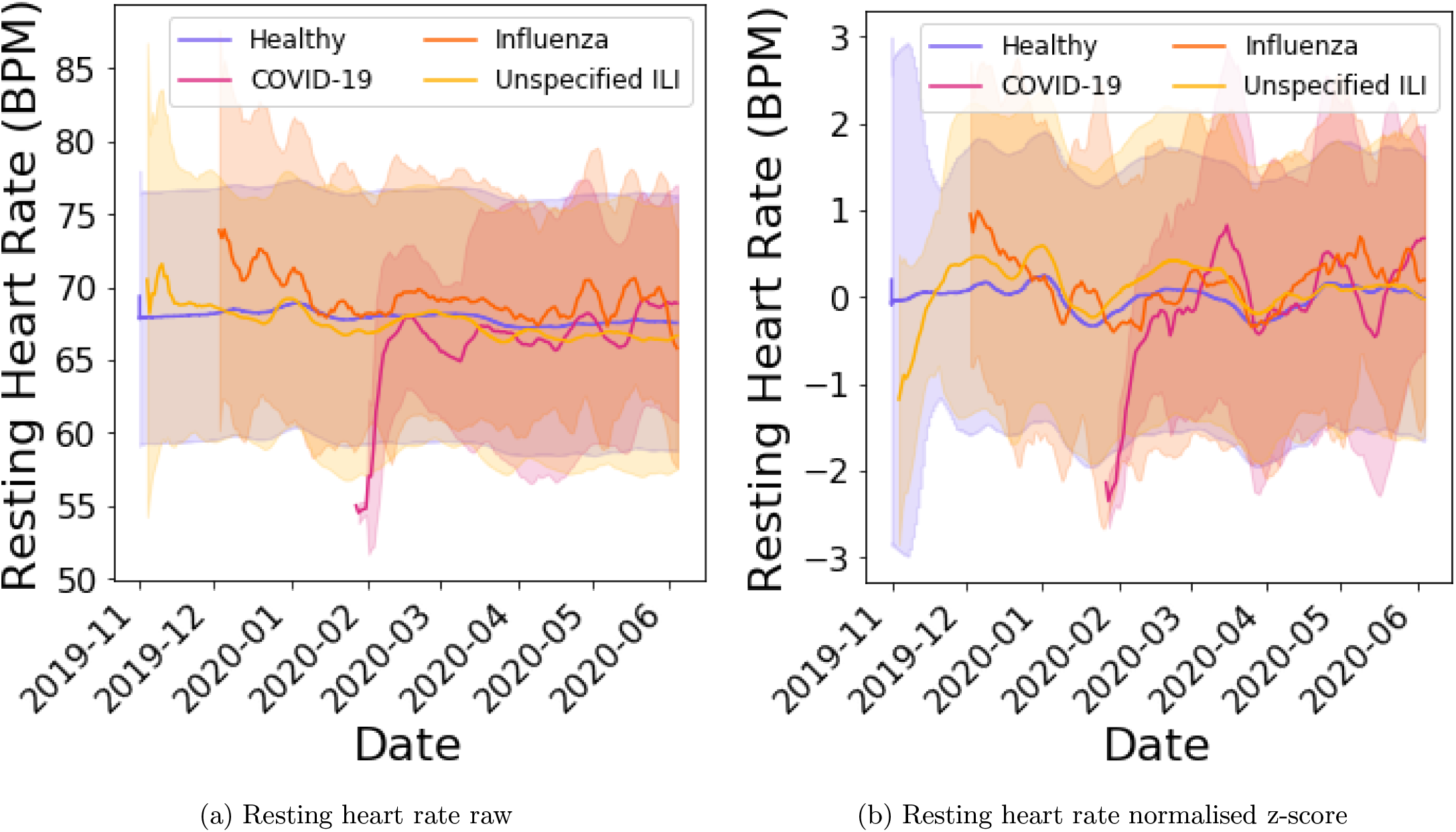
Heart rate feature throughout the study (mean ± standard deviation).

**Figure S4.**
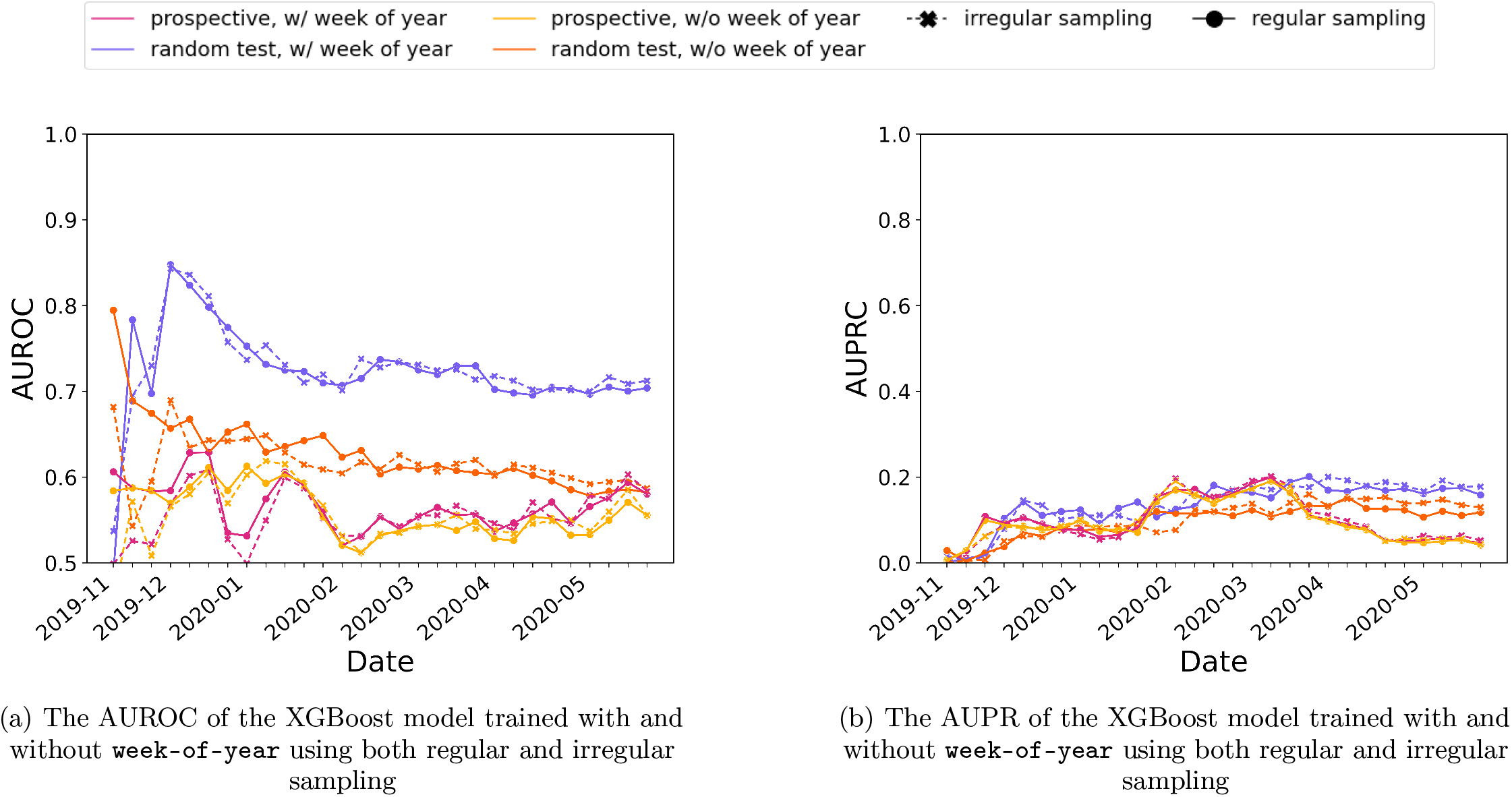
Regularly and irregularly sampled data trained on the same participant splits did not substantially affect the performance.

**Table S2.** Symptom prevalence among COVID negative and COVID positive ILI participants. Significance is tested for using a *χ*^2^ test with Bonferonni correction. Results with a *p* ≤ 0.01 are reject the null hypothesis and are shown in bold font.

| Feature | COVID-(%)<br>n=1389 | COVID+(%)<br>n=753 | *p* |
| --- | --- | --- | --- |
| Household member diagnosed with covid | 3.6% | 23.9% | < **0.0001** |
| Air Travel (14 days) | 8.1% | 29.2% | < **0.0001** |
| Contact ILI outside household | 16.9% | 36.3% | < **0.0001** |
| Contact covid | 10.0% | 35.6% | < **0.0001** |
| healthcare worker (household) | 3.0% | 2.4% | 0.0168 |
| healthcare worker (self) | 18.6% | 21.5% | **0.0031** |
| Anosmia | 11.2% | 19.5% | < **0.0001** |
| Chest pain | 20.7% | 31.1% | < **0.0001** |
| Short breath | 32.0% | 39.0% | < **0.0001** |
| Vaccinated this year (influenza) | 54.5% | 61.0% | **0.0007** |
| Fever | 25.5% | 29.7% | **0.0009** |
| No symptoms | 0.4% | 1.5% | < **0.0001** |
| Other | 3.2% | 1.3% | **0.0002** |
| Respiratory cough | 61.9% | 66.5% | **0.0049** |
| Respiratory nasal | 23.3% | 17.7% | < **0.0001** |
| Respiratory sneezing | 14.7% | 13.3% | 0.0143 |
| Respiratory sore throat | 28.7% | 19.3% | < **0.0001** |
| syst bodyache | 43.8% | 37.8% | **0.0006** |
| syst chills shiver | 21.2% | 21.6% | 0.0352 |
| syst fatigue | 28.9% | 24.0% | **0.0006** |
| syst headache | 39.2% | 29.9% | < **0.0001** |
| syst sweats | 18.0% | 18.7% | 0.0290 |

**Table S3.** Sparse data at irregular intervals

| Participant ID | Date | heart rate bpm | walk steps | sleep seconds |
| --- | --- | --- | --- | --- |
| 1 | 2020-03-11 | 71 | 4000 | *NaN* |
| 1 | 2020-03-14 | 74 | *NaN* | 28800 |
| 1 | 2020-03-15 | *NaN* | *NaN* | 26400 |
| 2 | 2020-02-21 | *NaN* | 5600 | 20100 |
| 2 | 2020-02-23 | 83 | *NaN* | *NaN* |

**Table S4.** Sparse data at regular intervals

| Participant ID | Date | heart rate bpm | walk steps | sleep seconds |
| --- | --- | --- | --- | --- |
| 1 | 2020-03-11 | 71 | 4000 | *NaN* |
| 1 | 2020-03-12 | *NaN* | *NaN* | *NaN* |
| 1 | 2020-03-13 | *NaN* | *NaN* | *NaN* |
| 1 | 2020-03-14 | 74 | *NaN* | 28800 |
| 1 | 2020-03-15 | *NaN* | *NaN* | 26400 |
| 2 | 2020-02-21 | *NaN* | 5600 | 20100 |
| 2 | 2020-02-22 | *NaN* | *NaN* | *NaN* |
| 2 | 2020-02-23 | 83 | *NaN* | *NaN* |

**Figure S5.**
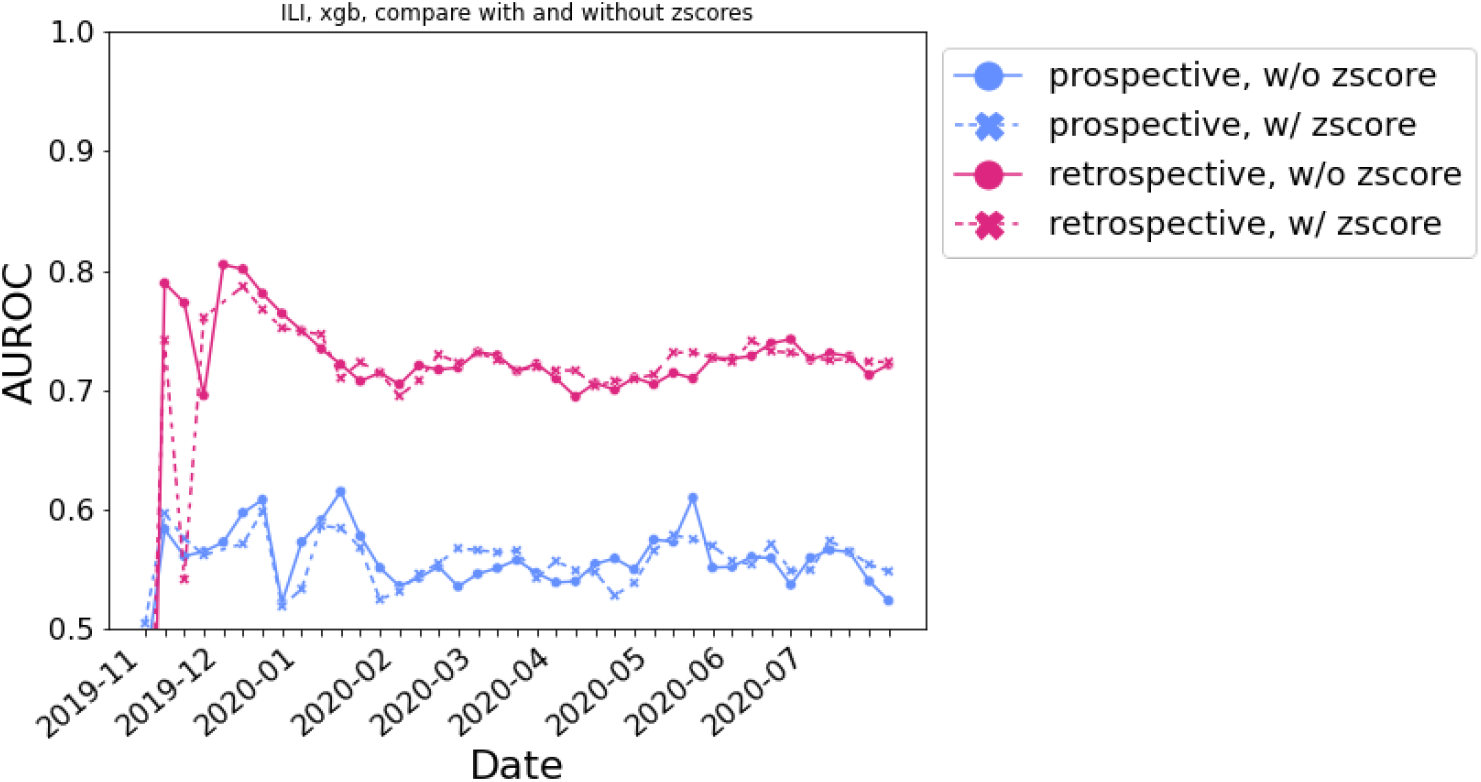
Comparison of z-score individually normalised features to population normalised features for XGBoost.

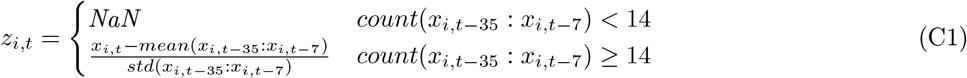

Where *x*_*i,t*_ is feature *i* at time *t*.

Otherwise, data are normalised by the mean features in the training data as shown in equation C2.

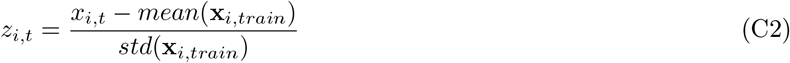

Where **x**_*i,train*_ is a vector of all observations of feature *i* in the training set.

In order for someone to have a feature observed on a certain day for the z-score normalisation, the raw feature must be observed *and* they must have at least 14 of the last 30 days of data as a source of standard deviation [51]. In contrast, when normalising data by the training population means, participants only need the single raw value in order for that day to be observed. We compare these two methods, referred to as z-score and standard data preparation in Figure S5. We do not see a large difference in performance, meaning the extra observations available in the standard data preparation regime offset any perceived gain of individual z-score normalisation. This is in spite of the inter-individual variance exceeding the intra-individual variance (See Figure S6b) as previously observed [35].

Next, data is imputed using *forward-filling*. At the earliest time-step for each participant, the training set population mean is applied (for z-score this corresponds to filling with 0), then the most recent measurement is carried forward in all subsequent observations. We retain the mask (whether or not the data was observed at that time-step), the time that has elapsed since the feature was last measured, and the forward-filled data [11]. The examples of regularly and irregularly sampled data are shown in Table S5 and Table S6, respectively. All pre-processing conditions (regularly vs. irregularly sampled data, imputed vs. non imputed, z-score vs raw data) are supported by the provided code.

### 1. Event reporting delay does not explain poor performance

The `FLUCOVID` dataset requires exclusively on self-reported events, including onset and recovery dates in order to form labels. Some efforts are taken to smooth events, like merging two ILI events for a participant that bookcase a healthy period of less than one week. We find that despite long delays in reporting illness, the sensitivity and specificity of the models on these labels are not substantially harmed (See Figure S7). Fortunately the majority of labels are reported within a 1-week period, and reporting lapses exponentially decay. However, all of our test sets include these participants in performance reporting nonetheless. Using outcomes attained directly from health systems may eliminate this perceived lapse in test performance and it could strengthen signals during training.

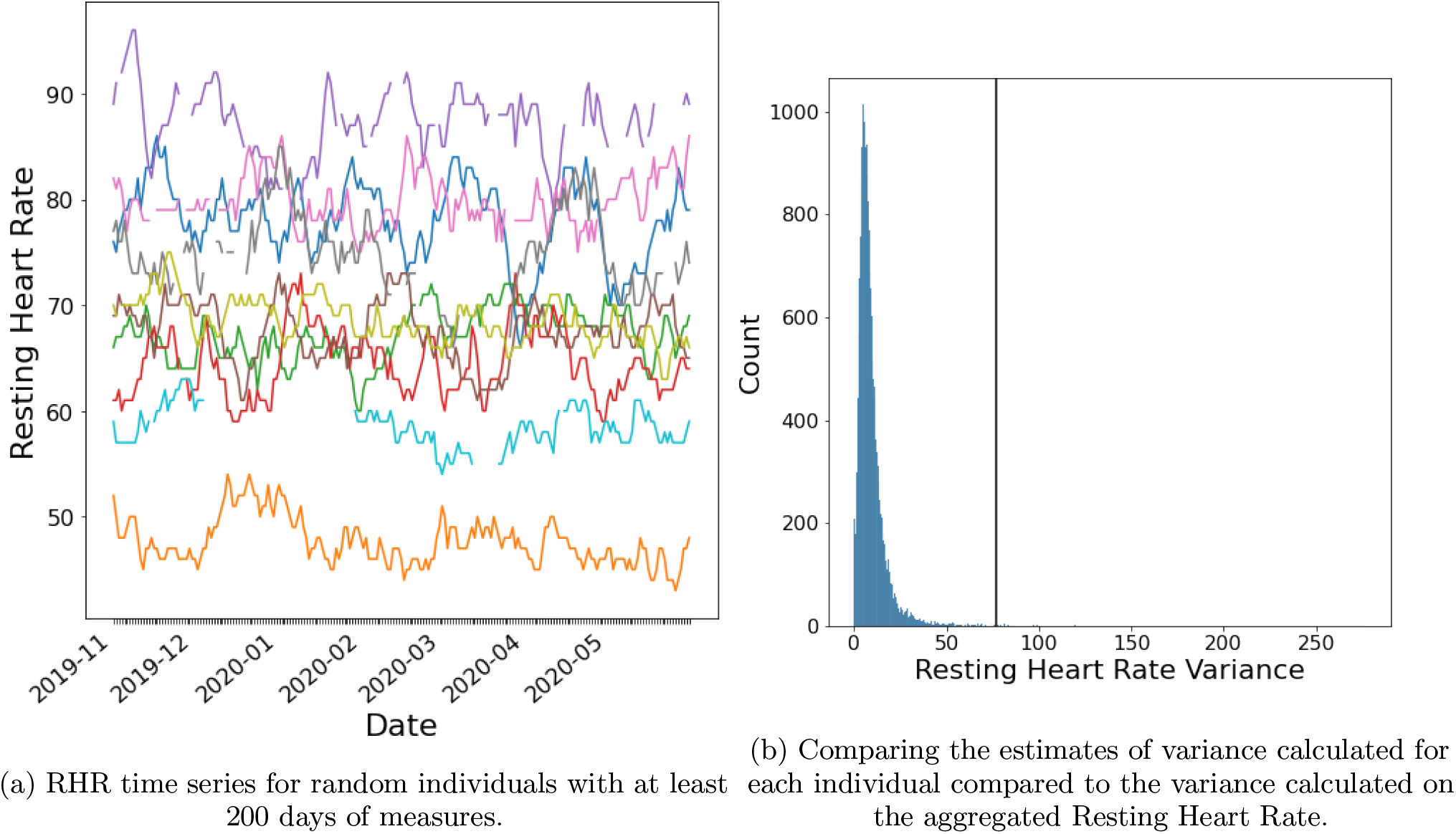

**Table S5.** Sparse data at regular intervals imputed using forward filling. The mask indicates if the data was observed at that time-point. The Δ*t* is the time since the last measurement was made. *Where data is not available at the start, it is imputed with the population mean.

| ID | Date | heart rate bpm |  |  | walk steps |  |  | sleep seconds |  |  |
| --- | --- | --- | --- | --- | --- | --- | --- | --- | --- | --- |
| | | *ffill* | mask | $\Delta t$ | *ffill* | mask | $\Delta t$ | *ffill* | mask | $\Delta t$ |
| 1 | 2020-03-11 | 71 | 1 | 0 | 4000 | 1 | 0 | 25100* | 0 | 0 |
| 1 | 2020-03-12 | 71 | 0 | 1 | 4000 | 0 | 1 | 25100* | 0 | 1 |
| 1 | 2020-03-13 | 71 | 0 | 2 | 4000 | 0 | 2 | 25100* | 0 | 2 |
| 1 | 2020-03-14 | 74 | 1 | 3 | 4000 | 0 | 3 | 28800 | 1 | 3 |
| 1 | 2020-03-15 | 74 | 0 | 1 | 4000 | 0 | 4 | 26400 | 1 | 1 |
| 2 | 2020-02-21 | 76* | 0 | 0 | 5600 | 1 | 0 | 20100 | 1 | 0 |
| 2 | 2020-02-22 | 76* | 0 | 1 | 5600 | 0 | 1 | 20100 | 0 | 1 |
| 2 | 2020-02-23 | 83 | 1 | 2 | 5600 | 0 | 2 | 20100 | 0 | 2 |

**Table S6.** Sparse data at irregular intervals imputed using forward filling. The mask indicates if the data was observed at that time-point. The Δ*t* is the time since the last measurement was made. *Where data is not available at the start, it is imputed with the population mean.

| ID | Date | heart rate bpm |  |  | walk steps |  |  | sleep seconds |  |  |
| --- | --- | --- | --- | --- | --- | --- | --- | --- | --- | --- |
| | | *ffill* | mask | $\Delta t$ | *ffill* | mask | $\Delta t$ | *ffill* | mask | $\Delta t$ |
| 1 | 2020-03-11 | 71 | 1 | 0 | 4000 | 1 | 0 | 25100* | 0 | 0 |
| 1 | 2020-03-14 | 74 | 1 | 3 | 4000 | 0 | 3 | 28800 | 1 | 3 |
| 1 | 2020-03-15 | 74 | 0 | 1 | 4000 | 0 | 4 | 26400 | 1 | 1 |
| 2 | 2020-02-21 | 76* | 0 | 0 | 5600 | 1 | 0 | 20100 | 1 | 0 |
| 2 | 2020-02-23 | 83 | 1 | 2 | 5600 | 0 | 2 | 20100 | 0 | 2 |

**Figure S7.**
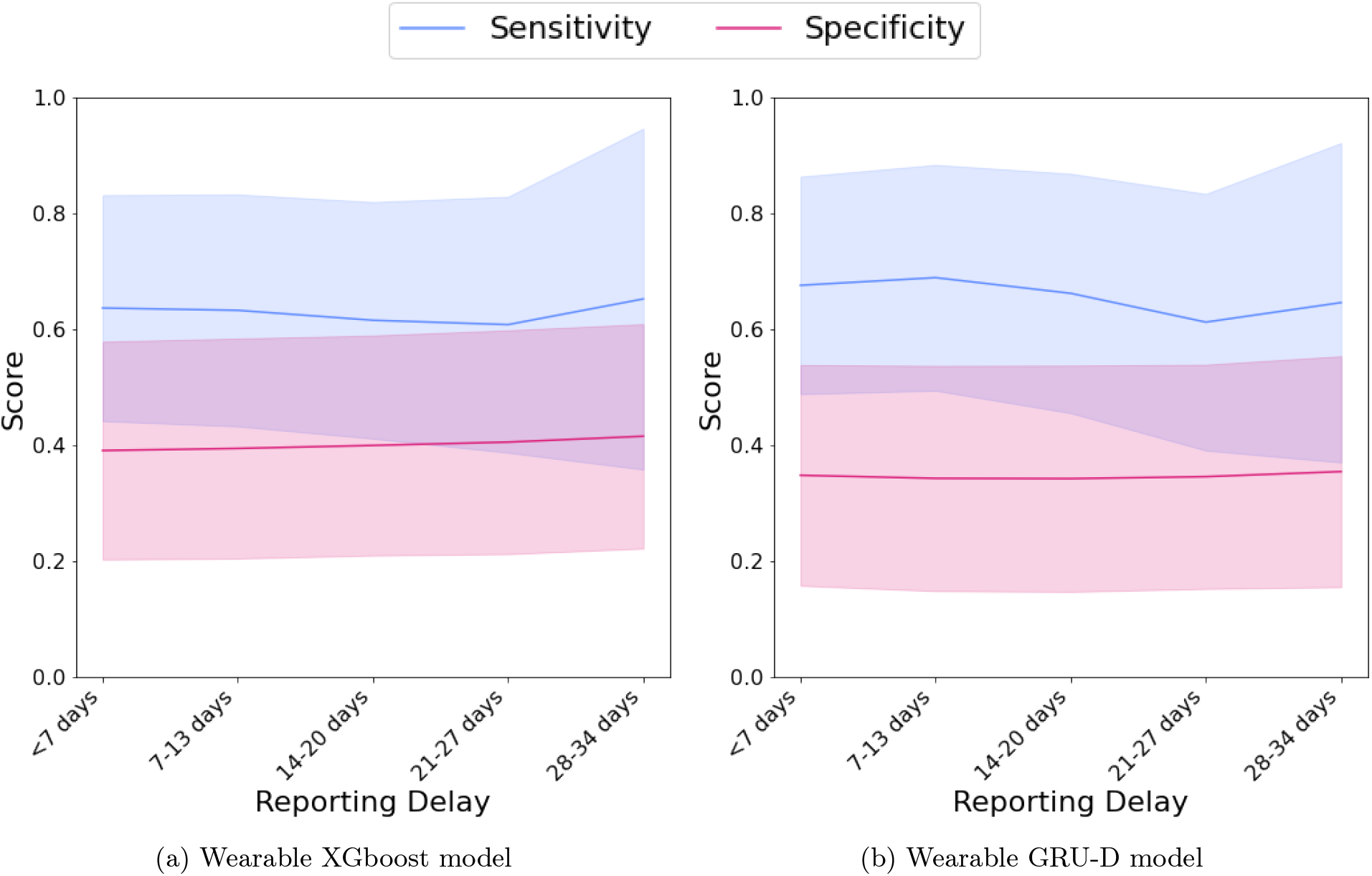
Performance of the ILI detection Wearable model, stratified by how far in the past an individual was remembering the event when responding to the survey

### 2. Prospective validation set matches prospective test set logits better than randomise validation set

## Appendix D: Comparison to existing models according to their evaluation schemes

We show performances of models as found in their respective citations in Table S7. Wherever possible we match their evaluation scheme for retrospective test sets, prospective test sets, and XGBoost and GRU-D models. Note, that we do not specifically optimise for these objectives, but instead simply select the appropriate days and calculate the metrics.

**Table S7.** Comparison of our models to literature with retrospective and prospective evaluations. *86 Controls. ** there was no significant difference between alarm rates for COVID-19 cases and potentially healthy cases, so the TPR*∼*FPR, or specificity*∼*0.125

| Paper | - Days | + Days | N | AUROC | AUPR | Sens. | Spec. | PPV | NPV |
| --- | --- | --- | --- | --- | --- | --- | --- | --- | --- |
| FitBit [43] | -21, -8 | 1, 8 | 196 | $0.77 \pm 0.02$ | - | $0.51 \pm 0.03$ | 0.9 | - | - |
| Ours (Random, XGBoost) | -21, -8 | 1, 8 | 204 | $0.59 \pm 0.11$ | $0.69 \pm 0.11$ | $0.12 \pm 0.13$ | $0.94 \pm 0.04$ | $0.64 \pm 0.28$ | $0.40 \pm 0.12$ |
| Ours (Random, GRU-D) | -21, -8 | 1, 8 | 204 | - | - | 0. | 0. | 0. | 0. |
| Ours (Prosp., XGBoost+survey) | -21, -8 | 1, 8 | 204 | - | - | $0.68 \pm 0.20$ | $0.92 \pm 0.02$ | $0.02 \pm 0.01$ | $1.00 \pm 0.00$ |
| Ours (Prosp., GRU-D+survey) | -21, -8 | 1, 8 | 204 | - | - | $0.68 \pm 0.20$ | $0.92 \pm 0.03$ | $0.02 \pm 0.01$ | $1.00 \pm 0.00$ |
| WHOOP [35] | -30, -14 | -2, 3 | 24 | - | - | 0.365 | 0.953 | 0.738 | 0.806 |
| Ours (Random, XGBoost) | -30, -14 | -2, 3 | 204 | $0.61 \pm 0.11$ | $0.70 \pm 0.12$ | $0.16 \pm 0.13$ | $0.97 \pm 0.02$ | $0.67 \pm 0.32$ | $0.40 \pm 0.10$ |
| Ours (Random, GRU-D) | -30, -14 | -2, 3 | 204 | - | - | 0. | 0. | 0. | - |
| Ours (Prosp., XGBoost+survey) | -30, -14 | -2, 3 | 204 | - | - | $0.57 \pm 0.27$ | $0.95 \pm 0.00$ | $0.02 \pm 0.01$ | $1.00 \pm 0.00$ |
| Ours (Prosp., GRU-D+survey) | -30, -14 | -2, 3 | 204 | - | - | $0.56 \pm 0.27$ | $0.95 \pm 0.00$ | $0.02 \pm 0.01$ | $1.00 \pm 0.00$ |
| CuSum [38] | None | -30, inf | 24* | - | - | 0.875 | 0.125** | - | - |

**Figure S8.**
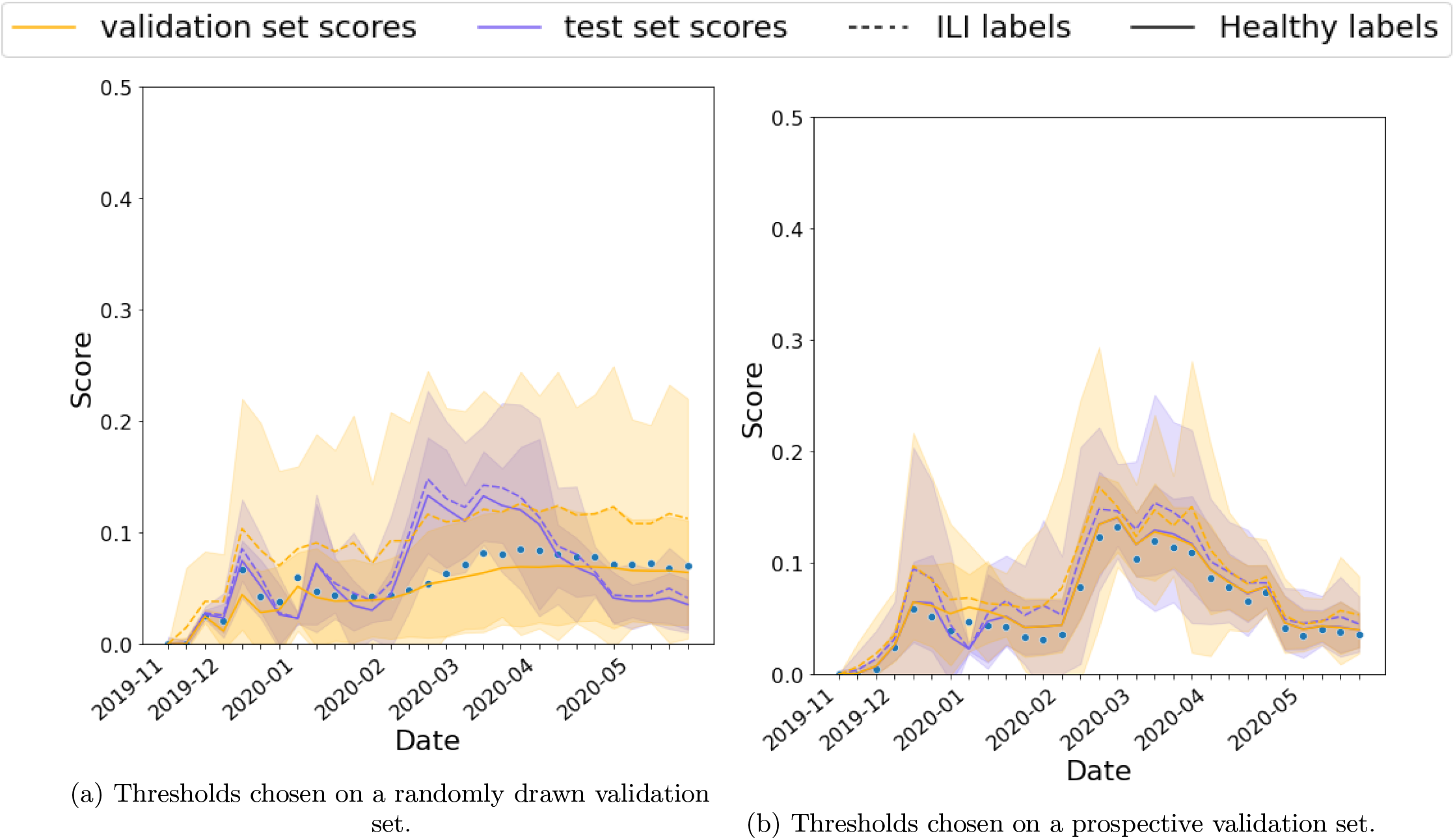
The distribution of the validation set must match the distribution of the test set when choosing thresholds.

**Table S8.**
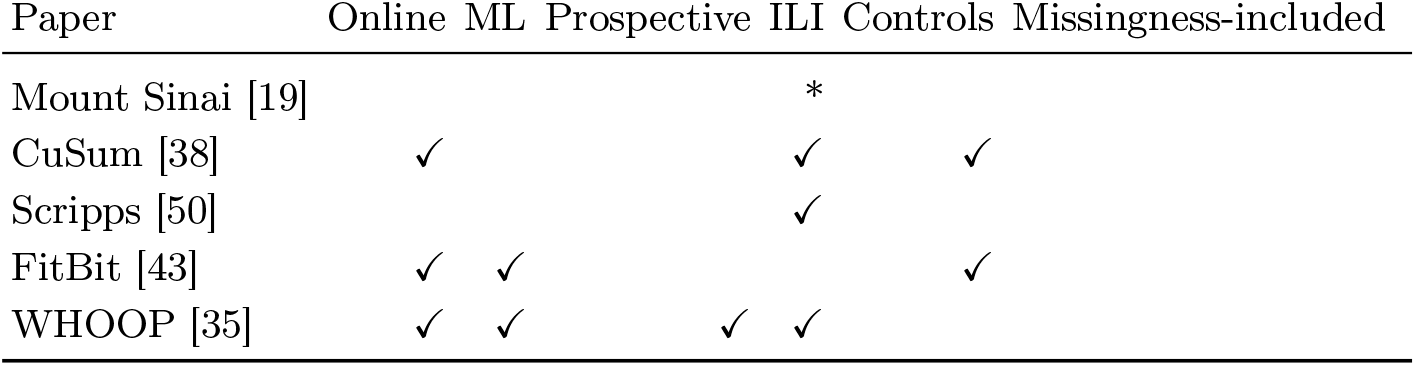
Methods in existing literature. None of the methods have evaluation schemes that accommodate participants with missing features. *For 13 patients, only 13 symptom days are compared to all other days. It is not clear how many of the symptom experiences overlapped with COVID-19 events.

## Appendix E: Time-to-onset plots of model performance

## Appendix F: Plots of model performance given different thresholds for missing data

We observed an effect on the performance of the models from missing data due to lack of device wear time. If an individual has not worn the device at all over a given week we see a drop in performance. This motivates work to improve the frequency which individuals in the study wear their devices.

**Figure S9.**
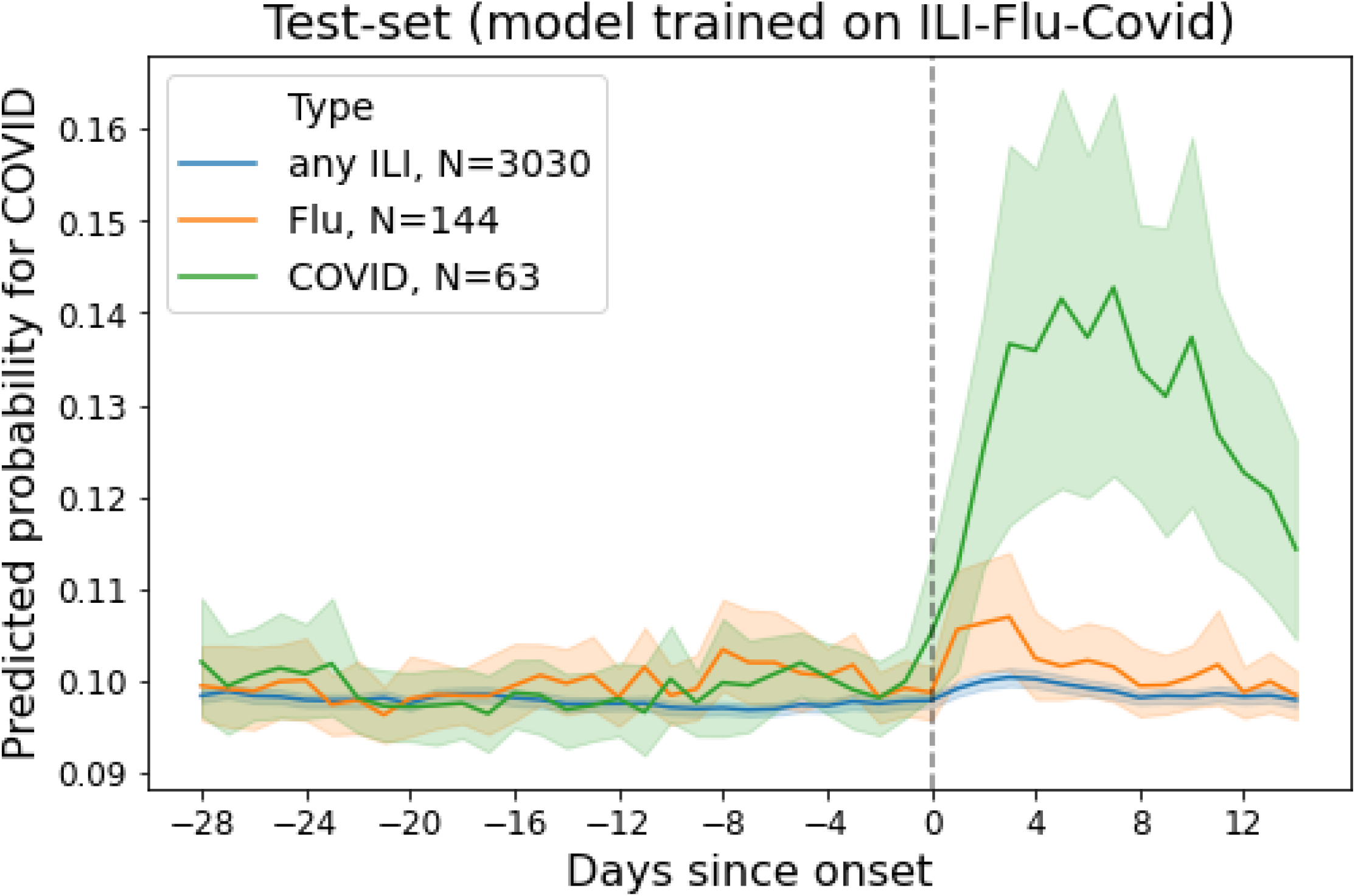
The wearable model outcome logits around symptom onset date for COVID-19, confirmed influenza, and unspecified ILI. The outcomes are gathered from the XGBoost model on retrospective, randomly selected test set.

**Figure S10.**
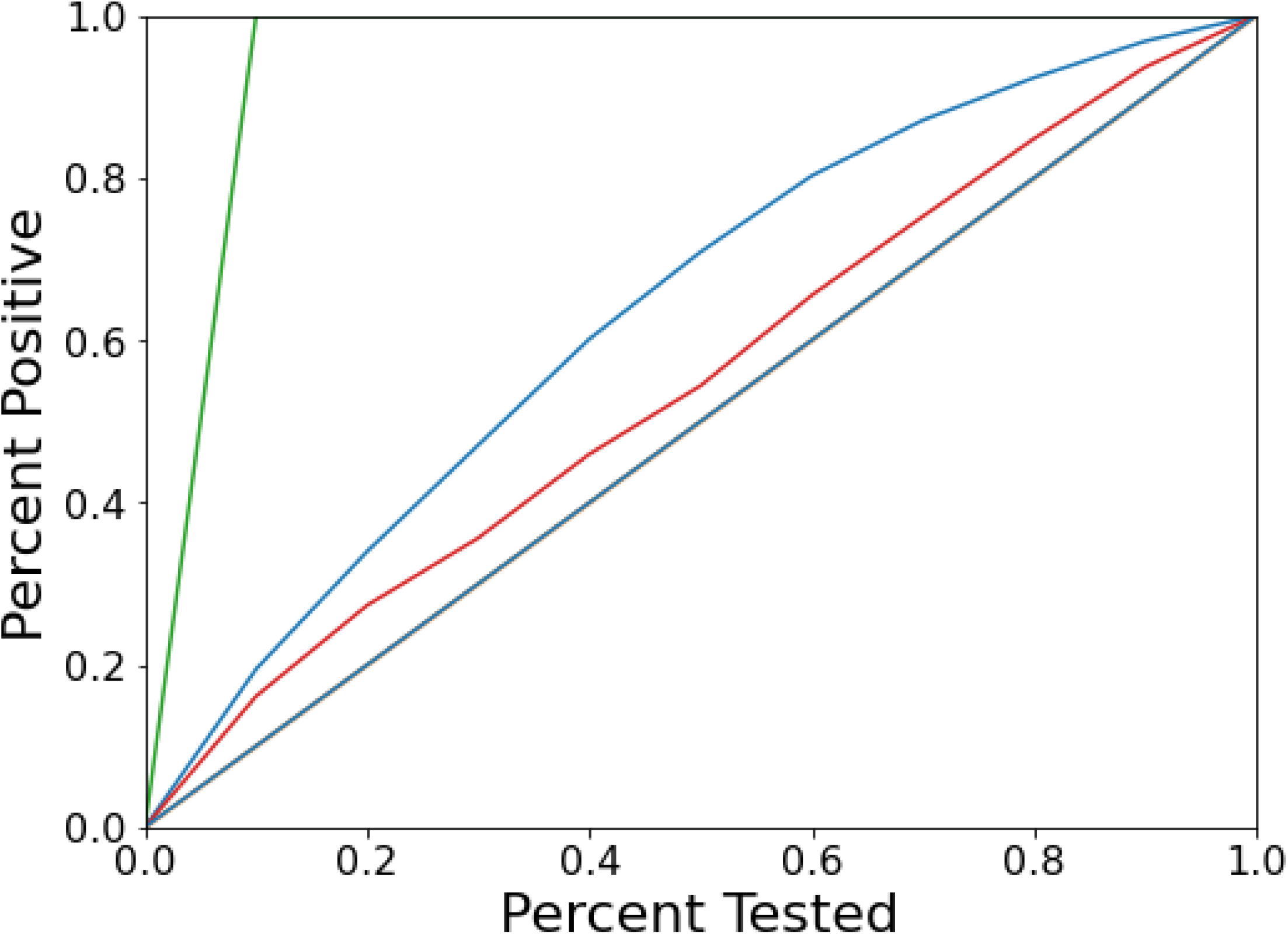
The gain of the model. Green represents the best-case scenario, blue is the model agnostic performance, and red is the when the data are normalised by the model.

**Figure S11.**
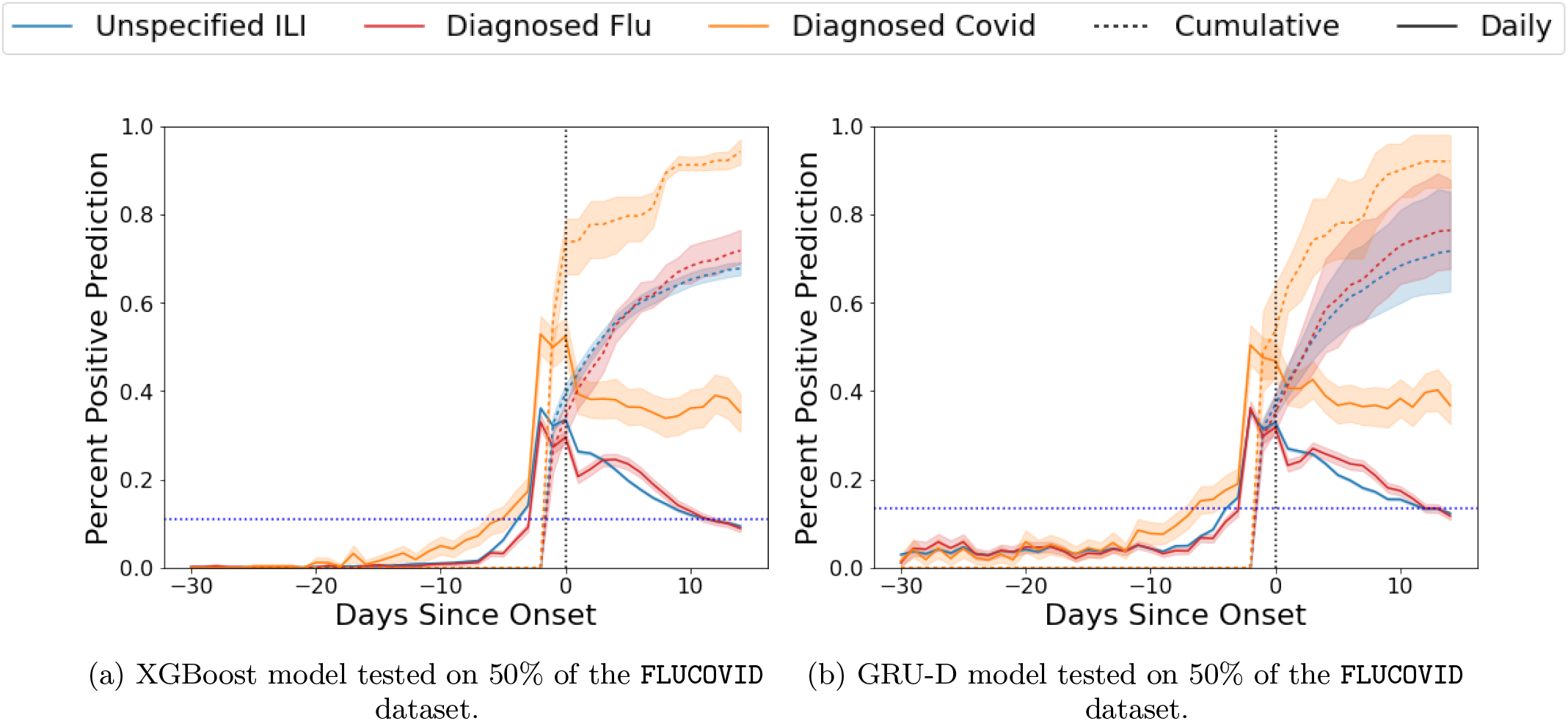
Performance of the Covid-19 Wearable+Survey detection models in the `FLUCOVID` dataset for both XGBoost (left) and GRU-D (right) wearable models aggregated by their time since onset. Cumulative predictions is for a subset of participants because of random sampling of participants each week.

**Figure S12.**
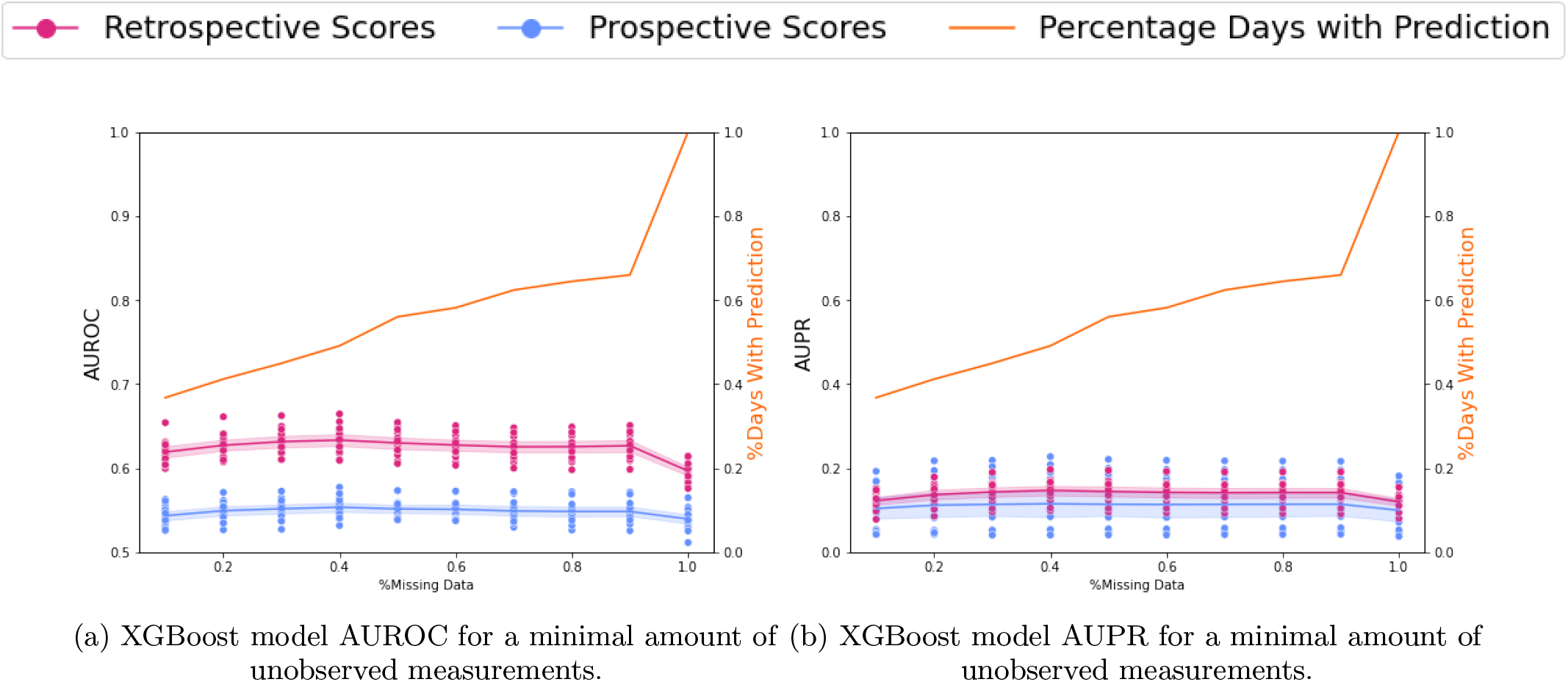
Varying the maximum amount of missing data allowed during testing affects the performance.

## Footnotes

1 Recent Emergency Use Authorization by FDA of rapid, off-the-counter at-home testing has yet to become widespread due to cost and logistics bottlenecks

## Notes

### Author Declarations

The study is approved by Solutions IRB (Initial approval date:, COVID-19 amendment: 4/3/2020)

## REFERENCES

[1] Adam, David (2020), “A guide to R-the pandemic’s misunderstood metric.” Nature 583 (7816), 346–348.

[2] Backer, Jantien A, Don Klinkenberg, and Jacco Wallinga (2020), “Incubation period of 2019 novel coronavirus (2019-nCoV) infections among travellers from wuhan, china, 20–28 january 2020,” Eurosurveillance 25 (5), 2000062.

[3] Bajema, Kristina L, Ryan E. Wiegand, Kendra Cuffe, Sadhna V. Patel, Ronaldo Iachan, Travis Lim, Adam Lee, Davia Moyse, Fiona P. Havers, Lee Harding, Alicia M. Fry, Aron J. Hall, Kelly Martin, Marjorie Biel, Yangyang Deng, III Meyer, William A., Mohit Mathur, Tonja Kyle, Adi V. Gundlapalli, Natalie J. Thornburg, Lyle R. Petersen, and Chris Edens (2020), “Estimated SARS-CoV-2 Seroprevalence in the US as of September 2020,” JAMA Internal Medicine 10.1001/jamainternmed.2020.7976, https://jamanetwork.com/journals/jamainternalmedicine/articlepdf/2773576/jamainternal_bajema_2020_oi_200107_1606151044.26955.pdf

[4] Bi, Qifang, Justin Lessler, Isabella Eckerle, Stephen A Lauer, Laurent Kaiser, Nicolas Vuilleumier, Derek AT Cummings, Antoine Flahault, Dusan Petrovic, Idris Guessous, Silvia Stringhini, and Andrew S Azman (2020), “Household transmission of SARS-COV-2: Insights from a population-based serological survey,” medRxiv 10.1101/2020.11.04.20225573, https://www.medrxiv.org/content/early/2020/11/04/2020.11.04.20225573.full.pdf.

[5] Byambasuren, Oyungerel, Magnolia Cardona, Katy Bell, Justin Clark, Mary-Louise McLaws, and Paul Glasziou (0), “Estimating the extent of asymptomatic COVID-19 and its potential for community transmission: Systematic review and meta-analysis,” Official Journal of the Association of Medical Microbiology and Infectious Disease Canada 0 (0), e20200030, https://doi.org/10.3138/jammi-2020-0030.

[6] Callahan, Alison, Ethan Steinberg, Jason A. Fries, Saurabh Gombar, Birju Patel, Conor K. Corbin, and Nigam H. Shah (2020), “Estimating the efficacy of symptom-based screening for COVID-19,” npj Digital Medicine 3 (1), 95.

[7] Carignan, Alex, Louis Valiquette, Cynthia Grenier, Jean Berchmans Musonera, Delphin Nkengurutse, Anaïs Marcil-Héguy, Kim Vettese, Dominique Marcoux, Corinne Valiquette, Wei Ting Xiong, Pierre-Hughes Fortier, Mélissa Généreux, and Jacques Pépin (2020), “Anosmia and dysgeusia associated with SARS-CoV-2 infection: an age-matched case–control study,” CMAJ 192 (26), E702–E707, https://www.cmaj.ca/content/192/26/E702.full.pdf.

[8] Centers for Disease Control and Prevention, (2020 (accessed September 18, 2020)), “CDC’s ILINet data,” https://gis.cdc.gov/grasp/fluview/fluportaldashboard.html.

[9] Cevik, Muge, Matthew Tate, Ollie Lloyd, Alberto Enrico Maraolo, Jenna Schafers, and Antonia Ho (XXXX), “SARS-CoV-2, SARS-CoV, and MERS-CoV viral load dynamics, duration of viral shedding, and infectiousness: a systematic review and meta-analysis,” The Lancet Microbe 10.1016/S2666-5247(20)30172-5.

[10] Challener, Douglas W, Gregory J. Challener, Vanessa J. Gow-Lee, Madiha Fida, Aditya S. Shah, and John C. O’Horo (2020), “Screening for COVID-19: Patient factors predicting positive pcr test,” Infection Control & Hospital Epidemiology 41 (8), 968–969.

[11] Che, Zhengping, Sanjay Purushotham, Kyunghyun Cho, David Sontag, and Yan Liu (2018), “Recurrent neural networks for multivariate time series with missing values,” Scientific reports 8 (1), 1–12.

[12] Chen, Tianqi, and Carlos Guestrin (2016), “XGBoost: A scalable tree boosting system,” in Proceedings of the 22nd ACM SIGKDD International Conference on Knowledge Discovery and Data Mining, KDD ’16 (ACM, New York, NY, USA) pp. 785–794.

[13] DeMasi, Orianna, Konrad Kording, and Benjamin Recht (2017), “Meaningless comparisons lead to false optimism in medical machine learning,” PLOS ONE 12 (9), 1–15.

[14] Estabragh, Zahra Raisi, and Mamas A Mamas (2013), “The cardiovascular manifestations of influenza: a systematic review,” International journal of cardiology 167 (6), 2397–2403.

[15] Griffith, Gareth J, Tim T. Morris, Matthew J. Tudball, Annie Herbert, Giulia Mancano, Lindsey Pike, Gemma C. Sharp, Jonathan Sterne, Tom M. Palmer, George Davey Smith, Kate Tilling, Luisa Zuccolo, Neil M. Davies, and Gibran Hemani (2020), “Collider bias undermines our understanding of covid-19 disease risk and severity,” Nature Communications 11 (1), 5749.

[16] Group, Delphi (2020 (accessed September 18, 2020)), “Covidcast,”.

[17] He, Xi, Eric H. Y. Lau, Peng Wu, Xilong Deng, Jian Wang, Xinxin Hao, Yiu Chung Lau, Jessica Y. Wong, Yujuan Guan, Xinghua Tan, Xiaoneng Mo, Yanqing Chen, Baolin Liao, Weilie Chen, Fengyu Hu, Qing Zhang, Mingqiu Zhong, Yanrong Wu, Lingzhai Zhao, Fuchun Zhang, Benjamin J. Cowling, Fang Li, and Gabriel M. Leung (2020), “Temporal dynamics in viral shedding and transmissibility of COVID-19,” Nature Medicine 26 (5), 672–675.

[18] Hicks, Jennifer L, Tim Althoff, Rok Sosic, Peter Kuhar, Bojan Bostjancic, Abby C. King, Jure Leskovec, and Scott L. Delp (2019), “Best practices for analyzing large-scale health data from wearables and smartphone apps,” npj Digital Medicine 2 (1), 45.

[19] Hirten, Robert P, Matteo Danieletto, Lewis Tomalin, Katie Hyewon Choi, Micol Zweig, Eddye Golden, Sparshdeep Kaur, Drew Helmus, Anthony Biello, Renata Pyzik, Alexander Charney, Riccardo Miotto, Benjamin S Glicksberg, Matthew Levin, Ismail Nabeel, Judith Aberg, David Reich, Dennis Charney, Erwin P Bottinger, Laurie Keefer, Mayte Suarez-Farinas, Girish N Nadkarni, and Zahi A Fayad (2021), “Use of physiological data from a wearable device to identify SARS-CoV-2 infection and symptoms and predict COVID-19 diagnosis: Observational study,” J Med Internet Res 23 (2), e26107.

[20] Hripcsak, George, David J Albers, and Adler Perotte (2015), “Parameterizing time in electronic health record studies,” Journal of the American Medical Informatics Association 22 (4), 794–804.

[21] Ip, Dennis K M, Lincoln L. H. Lau, Nancy H. L. Leung, Vicky J. Fang, Kwok-Hung Chan, Daniel K. W. Chu, Gabriel M. Leung, J. S. Malik Peiris, Timothy M. Uyeki, and Benjamin J. Cowling (2017), “Viral shedding and transmission potential of asymptomatic and paucisymptomatic influenza virus infections in the community,” Clinical infectious diseases : an official publication of the Infectious Diseases Society of America 64 (6), 736–742, 28011603[pmid].

[22] Jin, Jin, Neha Agarwala, Prosenjit Kundu, Benjamin Harvey, Yuqi Zhang, Eliza Wallace, and Nilanjan Chatterjee (2021), “Individual and community-level risk for COVID-19 mortality in the united states,” Nature Medicine 27 (2), 264–269.

[23] Kabarriti, Rafi, N. Patrik Brodin, Maxim I. Maron, Chandan Guha, Shalom Kalnicki, Madhur K. Garg, and Andrew D. Racine (2020), “Association of Race and Ethnicity With Comorbidities and Survival Among Patients With COVID-19 at an Urban Medical Center in New York,” JAMA Network Open 3 (9), e2019795–e2019795, https://jamanetwork.com/journals/jamanetworkopen/articlepdf/2770960/kabarriti_2020_oi_200689_1600295041.53638.pdf.

[24] Karjalainen, Jouko, and Matti Viitasalo (1986), “Fever and Cardiac Rhythm,” Archives of Internal Medicine 146 (6), 1169–1171, https://jamanetwork.com/journals/jamainternalmedicine/articlepdf/606966/archinte_146_6_026.pdf.

[25] Kavanagh, Matthew M, Ngozi A. Erondu, Oyewale Tomori, Victor J. Dzau, Emelda A. Okiro, Allan Maleche, Ifeyinwa C. Aniebo, Umunya Rugege, Charles B. Holmes, and Lawrence O. Gostin (2020), “Access to lifesaving medical resources for african countries: COVID-19 testing and response, ethics, and politics,” The Lancet 395 (10238), 1735–1738.

[26] Kleinman, Robert A, and Colin Merkel (2020), “Digital contact tracing for COVID-19,” CMAJ 192 (24), E653–E656, https://www.cmaj.ca/content/192/24/E653.full.pdf.

[27] Kucirka, Lauren M, Stephen A. Lauer, Oliver Laeyendecker, Denali Boon, and Justin Lessler (2020), “Variation in falsenegative rate of reverse transcriptase polymerase chain reaction–based SARS-CoV-2 tests by time since exposure,” Annals of Internal Medicine 173 (4), 262–267.

[28] Larremore, Daniel B, Bryan Wilder, Evan Lester, Soraya Shehata, James M. Burke, James A. Hay, Milind Tambe, Michael J. Mina, and Roy Parker (2020), “Test sensitivity is secondary to frequency and turnaround time for COVID-19 screening,” Science Advances 10.1126/sciadv.abd5393, https://advances.sciencemag.org/content/early/2020/11/20/sciadv.abd5393.1.full.pdf.

[29] Lauer, Stephen A, Kyra H. Grantz, Qifang Bi, Forrest K. Jones, Qulu Zheng, Hannah R. Meredith, Andrew S. Azman, Nicholas G. Reich, and Justin Lessler (2020), “The incubation period of coronavirus disease 2019 (COVID-19) from publicly reported confirmed cases: Estimation and application,” Annals of Internal Medicine 172 (9), 577–582.

[30] Lee, Seungjae, Tark Kim, Eunjung Lee, Cheolgu Lee, Hojung Kim, Heejeong Rhee, Se Yoon Park, Hyo-Ju Son, Shinae Yu, Jung Wan Park, Eun Ju Choo, Suyeon Park, Mark Loeb, and Tae Hyong Kim (2020), “Clinical Course and Molecular Viral Shedding Among Asymptomatic and Symptomatic Patients With SARS-CoV-2 Infection in a Community Treatment Center in the Republic of Korea,” JAMA Internal Medicine 180 (11), 1447–1452, https://jamanetwork.com/journals/jamainternalmedicine/articlepdf/2769235/jamainternal_lee_2020_oi_200057_1603479600.04745.pdf

[31] Lessler, Justin, Nicholas G. Reich, Ron Brookmeyer, Trish M. Perl, Kenrad E. Nelson, and Derek AT Cummings (2009), “Incubation periods of acute respiratory viral infections: a systematic review,” The Lancet Infectious Diseases 9 (5), 291–300.

[32] Leung, Nancy H L, Cuiling Xu, Dennis K. M. Ip, and Benjamin J. Cowling (2015), “Review article: The fraction of influenza virus infections that are asymptomatic: A systematic review and meta-analysis,” Epidemiology 26 (6).

[33] Li, Xiao, Jessilyn Dunn, Denis Salins, Gao Zhou, Wenyu Zhou, Sophia Miryam Schüssler-Fiorenza Rose, Dalia Perelman, Elizabeth Colbert, Ryan Runge, Shannon Rego, Ria Sonecha, Somalee Datta, Tracey McLaughlin, and Michael P. Snyder (2017), “Digital health: Tracking physiomes and activity using wearable biosensors reveals useful health-related information,” PLOS Biology 15 (1), 1–30.

[34] Menni, Cristina, Ana M. Valdes, Maxim B. Freidin, Carole H. Sudre, Long H. Nguyen, David A. Drew, Sajaysurya Ganesh, Thomas Varsavsky, M. Jorge Cardoso, Julia S. El-Sayed Moustafa, Alessia Visconti, Pirro Hysi, Ruth C. E. Bowyer, Massimo Mangino, Mario Falchi, Jonathan Wolf, Sebastien Ourselin, Andrew T. Chan, Claire J. Steves, and Tim D. Spector (2020), “Real-time tracking of self-reported symptoms to predict potential COVID-19,” Nature Medicine 26 (7), 1037–1040.

[35] Miller, Dean J, John V. Capodilupo, Michele Lastella, Charli Sargent, Gregory D. Roach, Victoria H. Lee, and Emily R. Capodilupo (2020), “Analyzing changes in respiratory rate to predict the risk of COVID-19 infection,” PLOS ONE 15 (12), 1–10.

[36] Mina, Michael J, Roy Parker, and Daniel B. Larremore (2020), “Rethinking covid-19 test sensitivity — a strategy for containment,” New England Journal of Medicine 383 (22), e120, https://doi.org/10.1056/NEJMp2025631.

[37] Mina, Michael J, Tim E. Peto, Marta García-Fiñana, Malcolm G. Semple, and Iain E. Buchan (2021), “Clarifying the evidence on sars-cov-2 antigen rapid tests in public health responses to covid-19,” The Lancet 10.1016/S0140-6736(21)00425-6.

[38] Mishra, Tejaswini, Meng Wang, Ahmed A. Metwally, Gireesh K. Bogu, Andrew W. Brooks, Amir Bahmani, Arash Alavi, Alessandra Celli, Emily Higgs, Orit Dagan-Rosenfeld, Bethany Fay, Susan Kirkpatrick, Ryan Kellogg, Michelle Gibson, Tao Wang, Erika M. Hunting, Petra Mamic, Ariel B. Ganz, Benjamin Rolnik, Xiao Li, and Michael P. Snyder (2020), “Pre-symptomatic detection of COVID-19 from smartwatch data,” Nature Biomedical Engineering 4 (12), 1208–1220.

[39] Mizumoto, Kenji, Katsushi Kagaya, Alexander Zarebski, and Gerardo Chowell (2020), “Estimating the asymptomatic proportion of coronavirus disease 2019 (COVID-19) cases on board the Diamond Princess cruise ship, Yokohama, Japan, 2020,” Eurosurveillance 25 (10), 2000180.

[40] Moreno-Torres, Jose G, Troy Raeder, Rocío Alaiz-Rodríguez, Nitesh V. Chawla, and Francisco Herrera (2012), “A unifying view on dataset shift in classification,” Pattern Recognition 45 (1), 521 – 530.

[41] Mozannar, Hussein, and David Sontag (2020), “Consistent estimators for learning to defer to an expert,” in Proceedings of the 37th International Conference on Machine Learning, Proceedings of Machine Learning Research, Vol. 119, edited by Hal Daumé Iii and Aarti Singh (PMLR) pp. 7076–7087.

[42] Muñoz-Price, L Silvia, Ann B. Nattinger, Frida Rivera, Ryan Hanson, Cameron G. Gmehlin, Adriana Perez, Siddhartha Singh, Blake W. Buchan, Nathan A. Ledeboer, and Liliana E. Pezzin (2020), “Racial Disparities in Incidence and Outcomes Among Patients With COVID-19,” JAMA Network Open 3 (9), e2021892–e2021892, https://jamanetwork.com/journals/jamanetworkopen/articlepdf/2770961/muozprice_2020_oi_200740_1600295049.06753.pdf.

[43] Natarajan, Aravind, Hao-Wei Su, and Conor Heneghan (2020), “Assessment of physiological signs associated with COVID-19 measured using wearable devices,” npj Digital Medicine 3 (1), 156.

[44] Paltiel, A David, Amy Zheng, and Rochelle P. Walensky (2020), “Assessment of SARS-CoV-2 Screening Strategies to Permit the Safe Reopening of College Campuses in the United States,” JAMA Network Open 3 (7), e2016818–e2016818, https://jamanetwork.com/journals/jamanetworkopen/articlepdf/2768923/paltiel_2020_oi_200614_1597251392.14363.pdf.

[45] Perez, Marco V, Kenneth W. Mahaffey, Haley Hedlin, John S. Rumsfeld, Ariadna Garcia, Todd Ferris, Vidhya Balasubramanian, Andrea M. Russo, Amol Rajmane, Lauren Cheung, Grace Hung, Justin Lee, Peter Kowey, Nisha Talati, Divya Nag, Santosh E. Gummidipundi, Alexis Beatty, Mellanie True Hills, Sumbul Desai, Christopher B. Granger, Manisha Desai, and Mintu P. Turakhia (2019), “Large-scale assessment of a smartwatch to identify atrial fibrillation,” New England Journal of Medicine 381 (20), 1909–1917, https://doi.org/10.1056/NEJMoa1901183.

[46] Pinker, Edieal (2018), “Reporting accuracy of rare event classifiers,” npj Digital Medicine 1 (1), 56.

[47] Pratap, Abhishek, Elias Chaibub Neto, Phil Snyder, Carl Stepnowsky, Noémie Elhadad, Daniel Grant, Matthew H. Mohebbi, Sean Mooney, Christine Suver, John Wilbanks, Lara Mangravite, Patrick J. Heagerty, Pat Areán, and Larsson Omberg (2020), “Indicators of retention in remote digital health studies: a cross-study evaluation of 100,000 participants,” npj Digital Medicine 3 (1), 21.

[48] Prem, Kiesha, Yang Liu, Timothy W. Russell, Adam J. Kucharski, Rosalind M. Eggo, Nicholas Davies, Stefan Flasche, Samuel Clifford, Carl A. B. Pearson, James D. Munday, Sam Abbott, Hamish Gibbs, Alicia Rosello, Billy J. Quilty, Thibaut Jombart, Fiona Sun, Charlie Diamond, Amy Gimma, Kevin van Zandvoort, Sebastian Funk, Christopher I. Jarvis, W. John Edmunds, Nikos I. Bosse, Joel Hellewell, Mark Jit, and Petra Klepac (2020), “The effect of control strategies to reduce social mixing on outcomes of the COVID-19 epidemic in wuhan, china: a modelling study,” The Lancet Public Health 5 (5), e261–e270.

[49] Price-Haywood, Eboni G, Jeffrey Burton, Daniel Fort, and Leonardo Seoane (2020), “Hospitalization and mortality among black patients and white patients with COVID-19,” New England Journal of Medicine 382 (26), 2534–2543, https://doi.org/10.1056/NEJMsa2011686.

[50] Quer, Giorgio, Jennifer M. Radin, Matteo Gadaleta, Katie Baca-Motes, Lauren Ariniello, Edward Ramos, Vik Kheterpal, Eric J. Topol, and Steven R. Steinhubl (2020), “Wearable sensor data and self-reported symptoms for COVID-19 detection,” Nature Medicine 10.1038/s41591-020-1123-x.

[51] Radin, Jennifer M, Nathan E. Wineinger, Eric J. Topol, and Steven R. Steinhubl (2020), “Harnessing wearable device data to improve state-level real-time surveillance of influenza-like illness in the usa: a population-based study,” The Lancet Digital Health 2 (2), e85–e93.

[52] Ren, Mengye, Wenyuan Zeng, Bin Yang, and Raquel Urtasun (2018), “Learning to reweight examples for robust deep learning,” in Proceedings of the 35th International Conference on Machine Learning, Proceedings of Machine Learning Research, Vol. 80, edited by Jennifer Dy and Andreas Krause (PMLR. Stockholmsmässsan, Stockholm Sweden) pp. 4334–4343.

[53] Shapiro, Allison, Nicole Marinsek, Ieuan Clay, Benjamin Bradshaw, Ernesto Ramirez, Jae Min, Andrew Trister, Yuedong Wang, Tim Althoff, and Luca Foschini (2021), “Characterizing covid-19 and influenza illnesses in the real world via person-generated health data,” Patterns 2 (1), 10.1016/j.patter.2020.100188.

[54] Sherman, Eli, Hitinder Gurm, Ulysses Balis, Scott Owens, and Jenna Wiens (2017), “Leveraging clinical time-series data for prediction: A cautionary tale,” AMIA Annu Symp Proc, 1571–1580.

[55] Smarr, Benjamin L, Kirstin Aschbacher, Sarah M. Fisher, Anoushka Chowdhary, Stephan Dilchert, Karena Puldon, Adam Rao, Frederick M. Hecht, and Ashley E. Mason (2020), “Feasibility of continuous fever monitoring using wearable devices,” Scientific Reports 10 (1), 21640.

[56] Spellberg, Brad, Travis B. Nielsen, and Arturo Casadevall (2020), “Antibodies, Immunity, and COVID-19,” JAMA Internal Medicine 10.1001/jamainternmed.2020.7986, https://jamanetwork.com/journals/jamainternalmedicine/articlepdf/2773575/jamainternal_spellberg_2020_ic_200042_1606151041.89445.pdf

[57] Surkova, Elena, Vladyslav Nikolayevskyy, and Francis Drobniewski (2020), “False-positive COVID-19 results: hidden problems and costs,” The Lancet Respiratory Medicine 8 (12), 1167–1168.

[58] Sze, Shirley, Daniel Pan, Clareece R. Nevill, Laura J. Gray, Christopher A. Martin, Joshua Nazareth, Jatinder S. Minhas, Pip Divall, Kamlesh Khunti, Keith R. Abrams, Laura B. Nellums, and Manish Pareek (2020), “Ethnicity and clinical outcomes in COVID-19: A systematic review and meta-analysis,” EClinicalMedicine 29, 10.1016/j.eclinm.2020.100630.

[59] Watson, Jessica, Penny F Whiting, and John E Brush (2020), “Interpreting a COVID-19 test result,” BMJ 369, 10.1136/bmj.m1808, https://www.bmj.com/content/369/bmj.m1808.full.pdf.

[60] Wölfel, Roman, Victor M. Corman, Wolfgang Guggemos, Michael Seilmaier, Sabine Zange, Marcel A. Müller, Daniela Niemeyer, Terry C. Jones, Patrick Vollmar, Camilla Rothe, Michael Hoelscher, Tobias Bleicker, Sebastian Brünink, Julia Schneider, Rosina Ehmann, Katrin Zwirglmaier, Christian Drosten, and Clemens Wendtner (2020), “Virological assessment of hospitalized patients with COVID-2019,” Nature 581 (7809), 465–469.

[61] Woloshin, Steven, Neeraj Patel, and Aaron S. Kesselheim (2020), “False negative tests for SARS-CoV-2 infection — challenges and implications,” New England Journal of Medicine 383 (6), e38. pMID: 32502334, https://doi.org/10.1056/NEJMp2015897.

[62] Xie, Zhigang, Ara Jo, and Young-Rock Hong (2020), “Electronic wearable device and physical activity among us adults: An analysis of 2019 hints data,” International Journal of Medical Informatics 144, 104297.

[63] Yan, Carol H, Farhoud Faraji, Divya P. Prajapati, Christine E. Boone, and Adam S. DeConde (2020), “Association of chemosensory dysfunction and COVID-19 in patients presenting with influenza-like symptoms,” International Forum of Allergy & Rhinology 10 (7), 806–813, https://onlinelibrary.wiley.com/doi/pdf/10.1002/alr.22579.

[64] Zhang, Lida, Nathan C. Hurley, Bassem Ibrahim, Erica Spatz, Harlan M. Krumholz, Roozbeh Jafari, and Mortazavi J. Bobak (2020), “Developing personalized models of blood pressure estimation from wearable sensors data using minimally-trained domain adversarial neural networks,” in Proceedings of the 5th Machine Learning for Healthcare Conference, Proceedings of Machine Learning Research, Vol. 126, edited by Finale Doshi-Velez, Jim Fackler, Ken Jung, David Kale, Rajesh Ranganath, Byron Wallace, and Jenna Wiens (PMLR, Virtual) pp. 97–120.

[65] Zhao, Qingyuan, Nianqiao Ju, Sergio Bacallado, and Rajen D. Shah (2020), “BETS: The dangers of selection bias in early analyses of the coronavirus disease (COVID-19) pandemic,” arXiv e-prints, 2004.07743 2004.07743 [stat.AP].

